# Wearable EEG during gameplay captures a robust P300 cognitive signal in unsupervised home settings

**DOI:** 10.64898/2026.05.10.26352556

**Authors:** Bernhard Specht, Andrej M. Savic, Samaher Garbaya, Reinhard Schneider, Djamel Khadraoui, Zied Tayeb

## Abstract

**Objective:** Continuous, unsupervised monitoring of cognitive brain responses has long been constrained by the demands of laboratory EEG. Whether the P300 event-related potential, an established marker of attention and cognitive processing, can be elicited as an incidental byproduct of genuine gameplay, recorded with a minimal wearable EEG system under unsupervised home conditions, has not been established.

**Approach:** Ten healthy adults played a gamified visual oddball task in which infrequent target stimuli (green gates) were embedded among frequent non-targets (red gates) within a continuous third-person running game. EEG was recorded with a four-channel dry-electrode headband (EEG channels: O1, O2, T3, T4; forehead reference; 250 Hz) with self-mounted electrodes in a home setting, without experimenter supervision. Group-level effects were assessed with cluster-based permutation tests and peak-amplitude tests. Single-trial classification used linear discriminant analysis (LDA) with four features per channel (16 total). Additional analyses included a within-subject comparison with a classical visual oddball paradigm using identical hardware, pilot data from a patient with relapsing–remitting multiple sclerosis, within-subject stability across 48 sessions, and pilot recordings with a headphone form factor.

**Main results:** A robust P300-like difference wave emerged on all four channels at the group level (cluster-based permutation tests, p < 0.05), with individual-level detection in 8 of 10 participants (exact binomial p <0.001). Single-trial LDA yielded a median cross-validated AUC of 0.730 (95% CI 0.672–0.820), with 9 of 10 participants exceeding chance. In a within-subject comparison, waveform morphology was closely preserved relative to a classical laboratory oddball, and classification performance was markedly higher in the game condition (AUC 0.820 versus 0.555). A patient with relapsing–remitting multiple sclerosis produced a clear P300 (AUC 0.853) with latencies within the healthy range. Within-session split-half reliability was high (r>0.70 on three of four channels), though between-session reliability was near zero across 48 sessions in one participant, with a declining classification trend over time. Pilot recordings with a headphone form factor also yielded a P300-like deflection.

**Significance:** These results demonstrate that the P300 can be elicited as a gameplay-integrated neural readout during genuine gameplay with a wearable, dry-electrode EEG system under unsupervised conditions. Gamification does not compromise P300 elicitation; in the within-subject comparison, it enhanced single-trial discriminability. The findings indicate that gamified, home-based P300 monitoring is achievable with minimal hardware and provide preliminary evidence for applicability in clinical populations, most notably multiple sclerosis, where P300 has established biomarker value but where the logistical burden of laboratory assessment currently precludes longitudinal use.

## 1 Introduction

Cognitive biomarkers derived from electroencephalography have an uncomfortable relationship with the clinic. The P300 event-related potential, elicited when an attended, infrequent stimulus is detected against a stream of frequent standards, is one of the most extensively validated markers of attentional resource allocation and context updating in the neuroscience literature (Polich, 2007). First described as a positive deflection occurring approximately 300 ms after an infrequent, task-relevant stimulus (Sutton et al., 1965; Picton, 1992), the P300 has become a standard marker of attentional resource allocation, stimulus evaluation, and context updating (Polich, 2007). Its latency prolongs, and its amplitude diminishes with cognitive decline (Polich, 2007), making it a sensitive index of disease burden in multiple sclerosis (Kaddoori, 2023; Vlieger et al., 2024) and migraine (Petrusic et al., 2022). Yet exploiting this sensitivity in practice requires a laboratory visit, a research-grade EEG system, and a willing experimenter. A single snapshot, obtained perhaps once per clinic appointment, is what passes for longitudinal monitoring.

A substantial body of work has explored P300 as a control signal for BCI-based spellers and menus (Fazel-Rezai et al., 2012; Finke et al., 2009; Congedo et al., 2011; Li et al., 2021). In these paradigms, the user intentionally attends to flashing targets to issue commands (Marshall et al., 2013; Kaplan et al., 2013). Systems such as the P300 speller have been adapted for gaming contexts with increasing sophistication, including the use of moving stimuli (Ganin et al., 2013). A recent systematic review of visual P300 BCI paradigms documents non-speller applications and confirms growing interest in wireless EEG hardware (Kalra et al., 2023). However, a fundamental constraint remains: in existing P300 gaming paradigms, the user’s cognitive task is to detect targets for BCI control, i.e. the P300 is the interface, not a byproduct of engagement with a meaningful game (Kaplan et al., 2013; Li et al., 2021).

Beyond fundamental research and BCIs, P300 has established clinical value as a cognitive biomarker in neurological populations. In multiple sclerosis (MS), P300 latency is prolonged and amplitude reduced relative to controls, with latency correlating positively with disease duration and EDSS (Kaddoori, 2023). A systematic review of ERP studies in MS confirms that P300 is a candidate cognitive biomarker, with visual stimuli yielding larger pooled effect sizes than auditory ones across ERP measures (Vlieger et al., 2024). Importantly, P300 amplitude and response speed relate not only to cognitive decline but also to preserved cognitive function in relapsing-remitting MS (Sundgren et al., 2015), raising the prospect that longitudinal P300 monitoring could track both deterioration and recovery. Similarly, P300 latency has been established as a biomarker for disease complexity in migraine with aura (Petrusic et al., 2022). Despite this clinical potential, current P300 assessment requires laboratory visits with standard EEG equipment, limiting the feasibility of frequent, longitudinal monitoring. A gamified, wearable approach that enables unsupervised P300 elicitation at home could address this gap, transforming cognitive monitoring from an episodic clinical measurement into a continuous, engaging, and scalable tool. Separately, efforts to validate P300 elicitation outside traditional laboratory settings have advanced along two fronts. First, virtual reality (VR) studies have shown that P300 can be reliably recorded with head-mounted displays (Kuziek et al., 2019), with P300-based BCI accuracy comparable to conventional monitors (Käthner et al., 2015), and that P300 can be elicited in immersive VR environments where the oddball requires no overt response (Chen et al., 2014). Second, the mobile brain/body imaging (MoBI) literature has demonstrated reliable P300 detection during real-world motor tasks such as walking, cycling, and driving (Grasso-Cladera et al., 2024), with auditory oddball paradigms remaining robust even in the presence of ecological background noise (Scanlon et al., 2019). These findings establish that P300 is not confined to static, controlled laboratory conditions, yet most ecological studies rely on auditory oddball paradigms overlaid on motor tasks, with visual oddball in genuinely interactive contexts remaining largely unexplored.

Advances in consumer-grade EEG hardware have further expanded the practical scope of P300 research. Comparative evaluations show that late ERP components, including P300, are detectable with consumer wireless devices, though temporal waveform fidelity may be reduced compared to research-grade systems (Lee et al., 2024). Miniaturized form factors such as around-the-ear flexible electrodes (Debener et al., 2015) have demonstrated viable P300 detection with non-standard electrode placements. However, such hardware validation studies have predominantly employed standard oddball tasks. Consumer-grade wearable EEG combined with a gamified, visually dynamic paradigm has not been tested, despite such combinations representing a natural convergence of wearable EEG and serious-game approaches for clinical monitoring.

Recent evidence suggests that game contexts may produce qualitatively different neural engagement than standard laboratory tasks. Pei et al. (2025) identified a P300-like component during gameplay characterized by unusually large magnitude and, critically, no habituation across repeated exposures. This finding stands in contrast to the well-documented amplitude reduction of P300 with stimulus repetition in conventional paradigms, and suggests that well-designed game mechanics may counteract neural adaptation, maintaining heightened cognitive engagement throughout the task. P300 amplitude has been shown to track individual preference for goal-score difficulty in a gamified educational task (Watanabe and Naruse, 2022), consistent with the notion that gamification may engage rather than disengage attentional processing of task-relevant stimuli.

These four lines of inquiry, P300-based BCI games, ecological validation, consumer-grade hardware, and gamification of neural engagement, have advanced independently, but no study sits at their intersection. Two additional gaps compound the problem: the consequences of forehead referencing for P300 polarity have not been characterized for wearable EEG devices, and longitudinal P300 stability in ecological paradigms is virtually unreported.

Despite these encouraging findings, gamifying the oddball paradigm poses a fundamental design challenge that has received little attention. The standard oddball task is, by construction, repetitive and monotonous: it requires a large number of frequent stimuli to establish the “standard” representation against which rare deviants are detected, and the deviant stimuli must be genuinely infrequent to elicit a robust P300. This means that recording sessions must be long enough to accumulate a sufficient number of deviant trials for reliable ERP averaging, while accounting for inevitable epoch rejections due to artifacts. These constraints are inherently at odds with engaging gameplay, which demands variety, unpredictability, and a fast pace. A further complication arises when the game includes motor control: in a game where the player actively moves (e.g., steering left or right), motor-related potentials can overlap with and contaminate the ERP of interest (Savić et al., 2014). If motor responses are systematically linked to stimulus category, the resulting movement-related activity becomes a confound that is inseparable from the cognitive P300 component. Thus, the central design problem is not merely whether P300 can be sustained in a game environment, but how to transform an inherently tedious, tightly constrained laboratory paradigm into an immersive, challenging, and enjoyable experience without violating the psychophysiological conditions required for robust P300 elicitation.

In this study, we present a gamified visual oddball task designed to address this challenge and evaluate the feasibility of P300 elicitation using a wearable dry-electrode EEG system. The task is a continuous third-person running game in which infrequent target stimuli (green gates) are embedded among frequent non-targets (red gates), preserving the statistical structure of a standard oddball paradigm within engaging gameplay. EEG was recorded using the headband, a four-channel dry-electrode device (O1, O2, T3, T4) with a forehead reference, while subjects played in a home setting with self-mounted electrodes. This configuration differs from standard P300 paradigms in three ways: (1) the oddball stimuli are integral to gameplay rather than serving as BCI control signals, (2) the visual environment is highly dynamic with continuous motion and complex backgrounds, and (3) the electrode montage and reference placement are non-standard, with no coverage of the canonical parietal P300 maximum.

We address the following specific aims. First, we test whether a P300-like difference wave emerges at the group and individual subject level despite the challenging recording conditions. Second, we evaluate single-trial classification performance to assess the practical detectability of the signal. Third, we present pilot data from a patient with multiple sclerosis to provide preliminary evidence of feasibility in a clinical population where P300 has an established biomarker value. We also report within-subject stability data across up to 48 repeated sessions, a within-subject comparison of the gamified and classical visual oddball paradigms using identical hardware, and pilot recordings from a headphone form factor of the same device (different channel positions).

Together, these results establish that gamified home-based P300 recording is not merely feasible but capable of producing signals of quality comparable to, and in some respects exceeding, those obtained in the laboratory.

## 2 Methods

### 2.1 Subjects

Ten healthy adults (8 male, 2 female; aged 28–37 years; all right-handed) with normal or corrected-to-normal vision and no history of neurological disease participated in the study (H1–H10). One patient with relapsing–remitting multiple sclerosis (P1) was additionally recruited as a clinical-population comparison. This study was approved by the Scientific Ethics Committee of the Clinical Center of Serbia and Neurology Clinic (reference number: 23-690). All standards and guidelines of the World Medical Association Declaration of Helsinki were respected. The participants signed a written informed consent form before participation.

### 2.2 Paradigms

#### 2.2.1 Classical visual oddball

A classical visual oddball task served as the control paradigm. The task was passive: participants fixated on a central cross and observed stimuli without producing a behavioural response. Each trial began with a fixation cross (duration jittered uniformly between 300 and 700 ms), followed by a coloured circle (red or yellow; figure 1) presented for 500 ms, and then a 1000 ms black screen. One colour was designated as the standard and the other as the deviant. Stimuli appeared in pseudo-random order with the constraint that at least two standards preceded each deviant. Each session comprised 300 trials (approximately 255 standard and 45 deviant; deviant probability ≈ 0.15) and lasted approximately 10 minutes. The empirical inter-stimulus onset interval had a median of 2.07 s.

**Figure 1:**
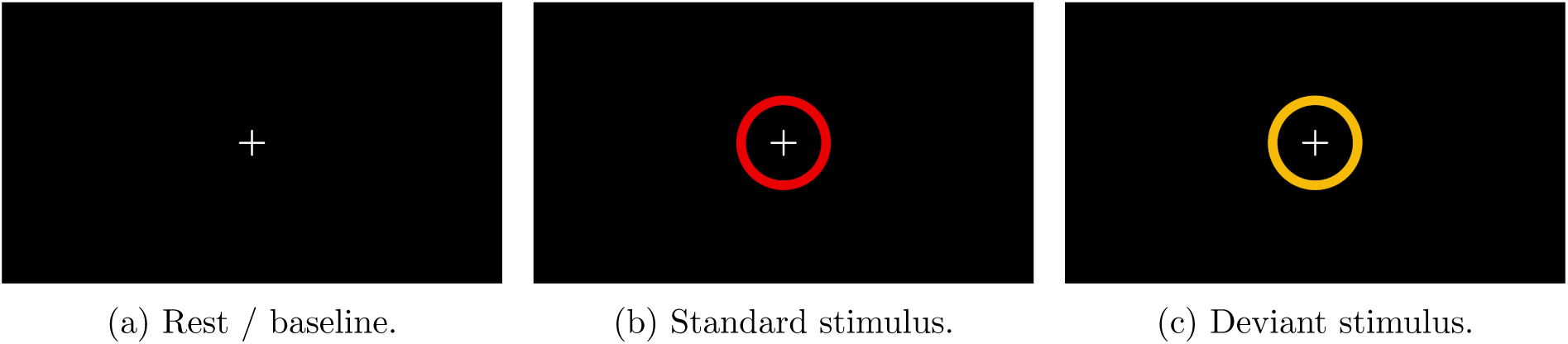
Screenshots from the classical visual oddball task.

#### 2.2.2 Gamified oddball

The gamified task was a continuous third-person running game implemented inside the Brain-Mirror app and played on a Samsung Galaxy Tab A8 tablet running Android 13 (figure 2). The player controlled an avatar running along a forest path with three lanes, swiping left or right to change lanes. Gates appeared ahead in the path, initially grey. As the avatar approached, each gate revealed its colour, as either red (standard) or green (deviant, target), the moment its in-game distance to the avatar fell below a fixed threshold (24 game units, corresponding to a median of 0.89 s of running before the avatar reached the gate plane). The grey-to-colour transition was designed to mirror the classical oddball structure: the grey gate served as the equivalent of the fixation cross, and the colour reveal as the stimulus onset.

**Figure 2:**
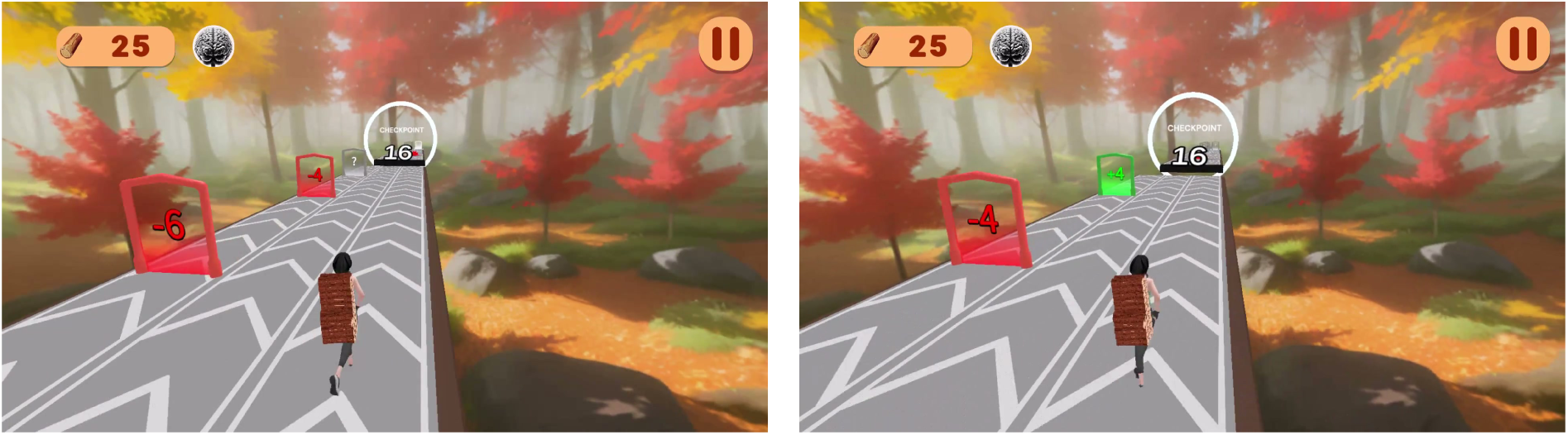
Gameplay screenshots from the gamified oddball task.

Passing through a green gate awarded points (displayed inside the gate); running into a red gate removed points. Points were translated into boards stacked on the avatar’s back, used to bridge gaps along the path. Gates were the only task-relevant stimuli, and the deviant category (green) was rare by design (approximately 1:5 deviant-to-standard ratio; cohort total 417 deviants vs. 2103 standards across 10 sessions). Sessions lasted a median of 7.3 min (range 4.9–7.9 min).

The avatar’s running speed was fixed, and the time between colour reveal and the avatar reaching the gate was jittered between 0.83 and 1.00 s (median 0.89 s; measured across all green-gate trials in the cohort), ensuring that motor responses (lane changes) were not time-locked to stimulus onset. The P3 response window (250–400 ms after colour reveal) therefore preceded the motor response, making the two temporally separable. Gate positions were pseudo-randomised across the three lanes, so that any movement-related potentials were distributed equally across conditions.

### 2.3 EEG recording

#### 2.3.1 Headband

The primary recordings used the headband, a consumer-grade wireless EEG device. The headband houses four gold-plated spring-loaded dry electrodes at positions O1, O2, T3, and T4 (international 10–20 system), referenced to a flat gold-plated forehead electrode; a separate forehead electrode serves as the common (ground) (figure 3). The device transmits data via Bluetooth Low Energy (BLE) at a sampling rate of 250 Hz with an input range of ±0.4 V and a signal bandwidth of 0–100 Hz. Battery life is up to 12 hours of continuous recording. The device holds FCC and CE certifications. Subjects self-mounted the headband following a graphical instruction manual provided by the custom graphical user interface, with impedance verification before each session. The custom interface displayed a per-channel contact-quality score on a 0–100 scale, derived from the electrode resistance (linearly mapped from 200 kΩ or below = 100 to 5 MΩ or above = 0), and colour-coded as red (quality < 52), yellow (52–82), or green (≥ 83). Subjects were instructed to reach at least the yellow state on every channel before starting a session, with green being the target. Self-mounting typically completed in under one minute, and no sessions were repeated due to poor contact. All recordings were performed in a home setting without experimenter supervision. The forehead reference has consequences for the observed P300 polarity. To produce waveforms visually consistent with the conventional P300 morphology, the difference wave throughout this paper is computed as standard minus deviant rather than the conventional deviant minus standard. This convention and its implications are discussed in detail in section 4.3.

**Figure 3:**
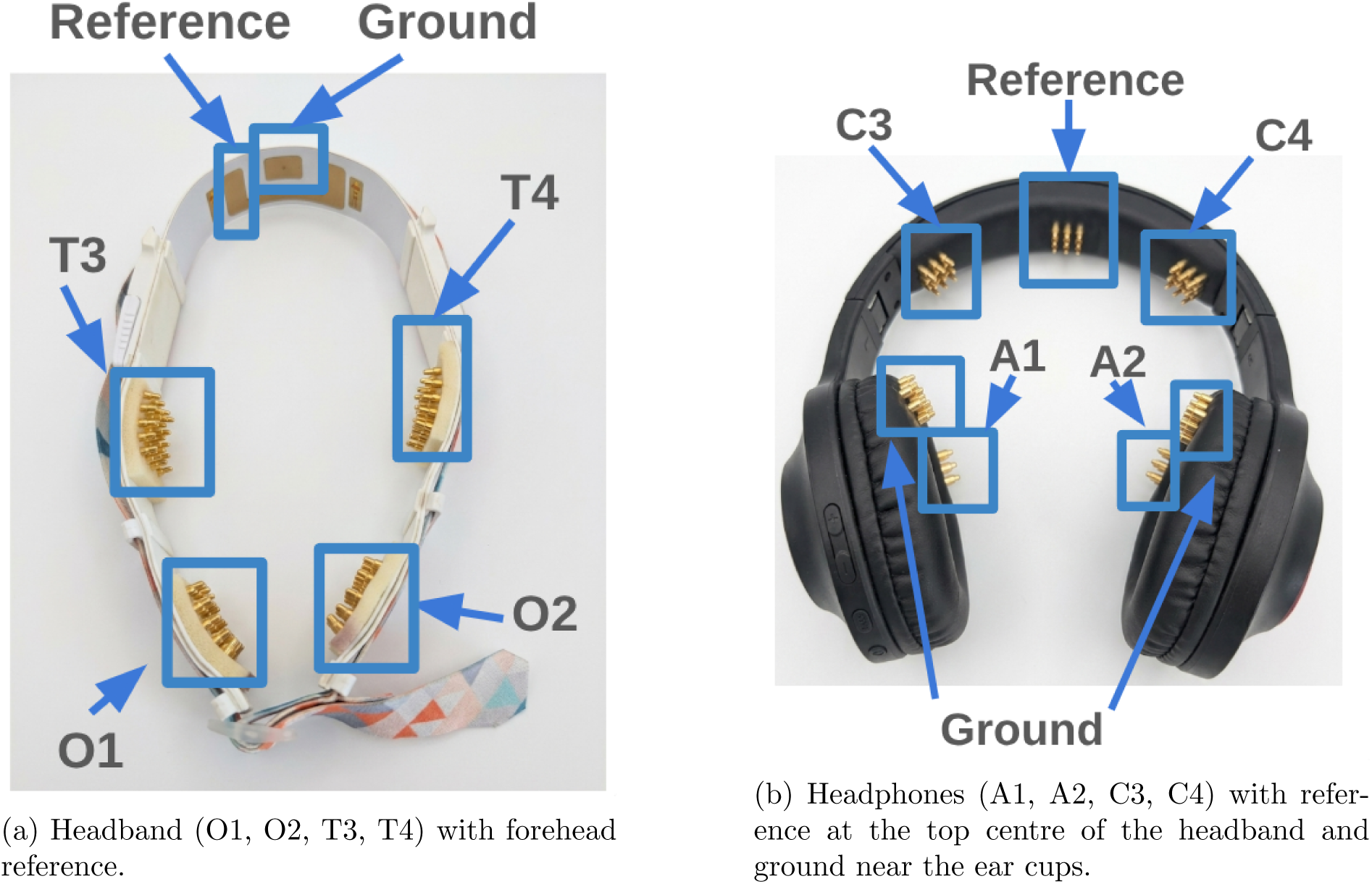
The electrode placement for the two form factors used in this study.

#### 2.3.2 Headphones

Two additional sessions were recorded using an EEG sensor (headphones), which places four gold-plated electrodes at A1, A2, C3, and C4 (figure 3b), with the reference electrode at the top centre of the headband (between C3 and C4, approximately at Cz) and the ground electrode at the bottom of the headband near the ear cups. Sampling rate and signal specifications are identical to the headband. These sessions are analysed separately (section 3.6).

### 2.4 Experimental design

Table 1 summarises the paradigms, hardware, and session counts per subject. For the primary group-level analyses, each healthy subject is represented by a single game session (their first). Subjects H1 and H2 contributed additional game sessions, reported as supplementary within-subject stability data (section 3.5). Subject H10 participated in an extended longitudinal recording of 48 game sessions and additionally completed 6 sessions of the classical visual oddball and 2 sessions with the headphone form factor, each analysed separately. Patient P1 completed a single game session on the headband hardware.

**Table 1:** Experimental design: paradigms, hardware, and session counts per subject.

| Subject(s) | Paradigm | Hardware | Sessions |
| --- | --- | --- | --- |
| H1–H10 | Gamified oddball | Headband | 1 each (primary analysis) |
| H1 | Gamified oddball | Headband | 4 additional (stability) |
| H2 | Gamified oddball | Headband | 2 additional (stability) |
| H10 | Gamified oddball | Headband | 47 additional (longitudinal) |
| H10 | Classical oddball | Headband | 6 |
| H10 | Gamified oddball | Headphones | 2 |
| P1 | Gamified oddball | Headband | 1 |

### 2.5 Preprocessing and epoching

Continuous EEG was band-pass filtered between 1 and 20 Hz and notch filtered at 50 Hz. Epochs were extracted from −200 to +700 ms relative to stimulus onset, baseline-corrected using the pre-stimulus interval (−200 to 0 ms), and rejected if the signal on any channel exceeded ±150 µV. For each subject, one standard, one deviant, and one difference (standard minus deviant) ERP was computed per channel by averaging the retained epochs.

### 2.6 Statistical analysis

All group-level tests treat the subject as the unit of observation. Normality was assessed with Shapiro–Wilk tests; parametric tests (one-sample or paired t-tests) were used when normality was not rejected, and Wilcoxon signed-rank tests otherwise. Tests across the four channels were corrected with the Benjamini–Hochberg false discovery rate (FDR).

#### Cluster-based permutation test

The temporal extent of the effect was quantified with a one-sample cluster-based permutation test on the per-subject difference waves (0–700 ms; 5000 permutations, two-sided).

#### Peak amplitude and paired tests

In the 250–400 ms window, the per-subject peak amplitude of the difference wave was tested against zero (one-sample), and the per-subject mean amplitude of the deviant evoked was compared to the standard evoked (paired). Effect sizes are reported as Cohen’s d or the rank-biserial correlation; Bayes factors (BF_10_, JZS prior) accompany the primary peak-amplitude test.

#### Single-subject detection

A one-sided Welch’s t-test on epoch-level mean amplitude in 250–400 ms (deviant < standard) was applied per subject, FDR-corrected across channels. Detection rates were tested against 5% chance using an exact binomial test.

#### Trial-level classification

A linear discriminant analysis (LDA) classifier (unshrunk, raw-µV features, no per-feature standardisation) was trained per subject using 5-fold stratified cross-validation with four per-channel features in the 250–400 ms window: mean amplitude, peak-to-peak amplitude, signed area under the curve, and zero-crossing count (16 features total). The feature set was selected *a priori* from standard ERP feature families and was not tuned on the present dataset. Per-subject AUC p-values were obtained from 1000 permutations of the class label vector (FDR-corrected across subjects). The group-level median AUC was tested against 0.5 with a two-sided Wilcoxon signed-rank test (10 000-resample bootstrap CI on the median).

#### Exploratory analyses

Spearman rank correlations (bootstrap 95% CI, 2000 resamples) were computed between task accuracy and both peak amplitude and LDA AUC. Within-session habituation was tested by splitting each subject’s epochs in half by trial order and comparing peak amplitudes.

## 3 Results

### 3.1 Group-level P300 and single-subject detection

The grand-average difference wave (standard minus deviant) across the ten healthy subjects showed a positive deflection in the 250–400 ms window on all four channels, consistent with a P300 response to the deviant (green) gate (figure 4). The effect was largest at occipital sites and smaller at temporal sites. Cluster-based permutation tests identified significant clusters covering the P3 window on all four channels (p < 0.05, 5000 permutations; significant windows are shaded in figure 4).

**Figure 4:**
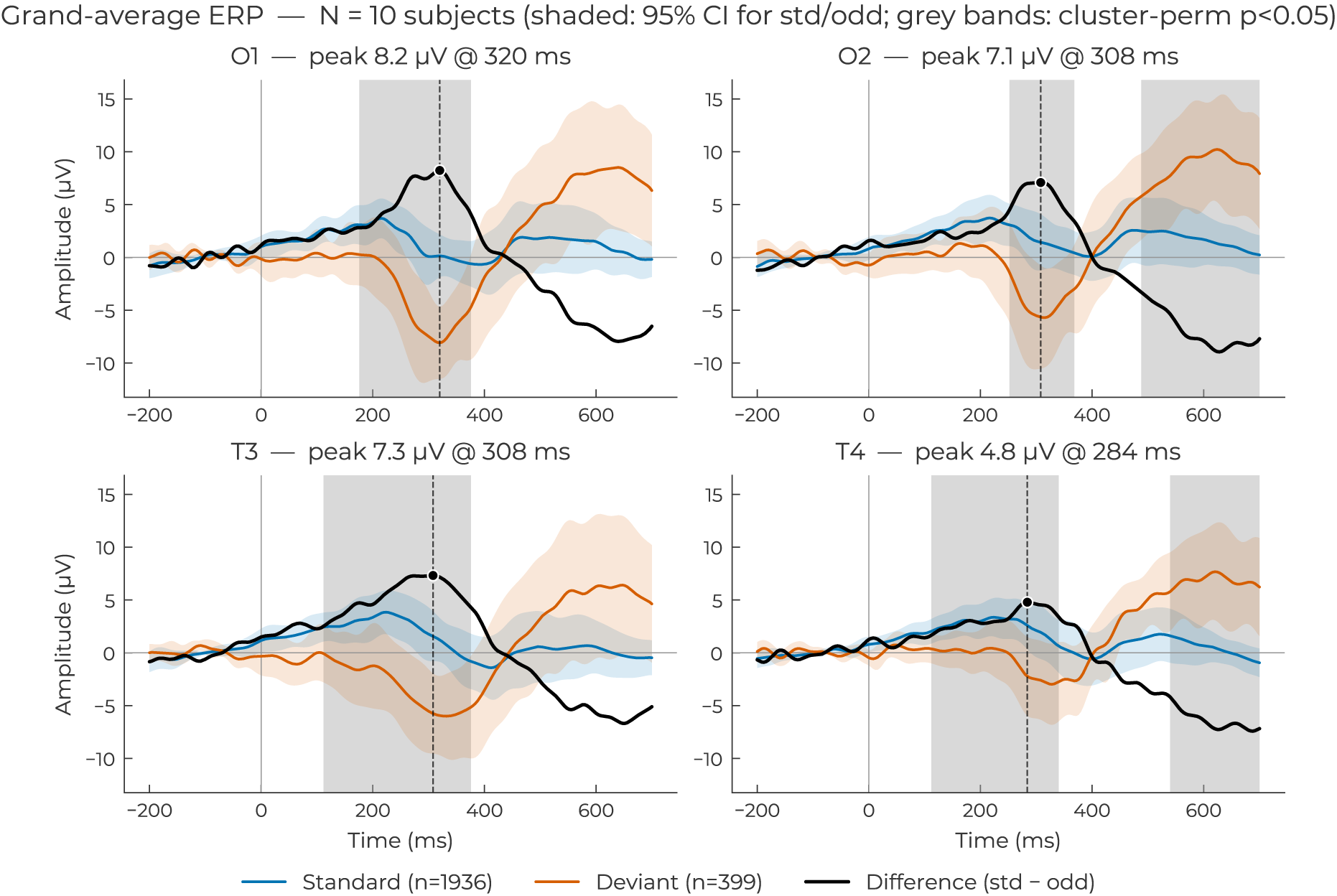
Grand-average ERP per channel (N = 10, equal weight per subject). Blue: standard; red: deviant; black: difference wave (standard − deviant). Shaded bands: 95% CIs. Yellow band: 250–400 ms analysis window. Grey shading indicates significant cluster windows from the permutation test.

Mean per-subject peak amplitudes in the 250–400 ms window were 11.76 µV (O1), 10.47 µV (O2), 8.73 µV (T3), and 6.09 µV (T4), with latencies tightly clustered between 322 and 338 ms across channels (supplementary figure S2). Effect sizes were very large on all channels (Cohen’s d = 3.84 [O1], 2.13 [O2], 1.41 [T3], 1.14 [T4]) and Bayes factors provided decisive evidence for a non-zero effect (BF_10_ = 2.1 × 10^4^ [O1], 329 [O2], 27.4 [T3], 9.8 [T4]). Diff-wave signal-to-noise ratios (peak amplitude divided by the pre-stimulus baseline standard deviation) had subject-level medians of 5.9 (O1), 7.6 (O2), 6.1 (T3), and 5.8 (T4). One-sample tests of the peak amplitude against zero were significant on all channels (all p_FDR_ ≤ 0.027). Paired comparisons of the deviant and standard ERPs in the same window confirmed the separation on all four channels (all p_FDR_ ≤ 0.006). Across the cohort, 7.3% of epochs were rejected by the ±150 µV threshold (per-subject median 7.5%, range 1.1–14.0%); per-subject final epoch counts ranged from 121–252 standards and 34–46 deviants (table 2). The per-subject difference waves (figure 5) show that the deflection was present in a majority of subjects, though amplitude varied substantially across individuals (see also the per-subject spaghetti plot in supplementary figure S1 and individual subject waveforms in supplementary figure S13).

**Figure 5:**
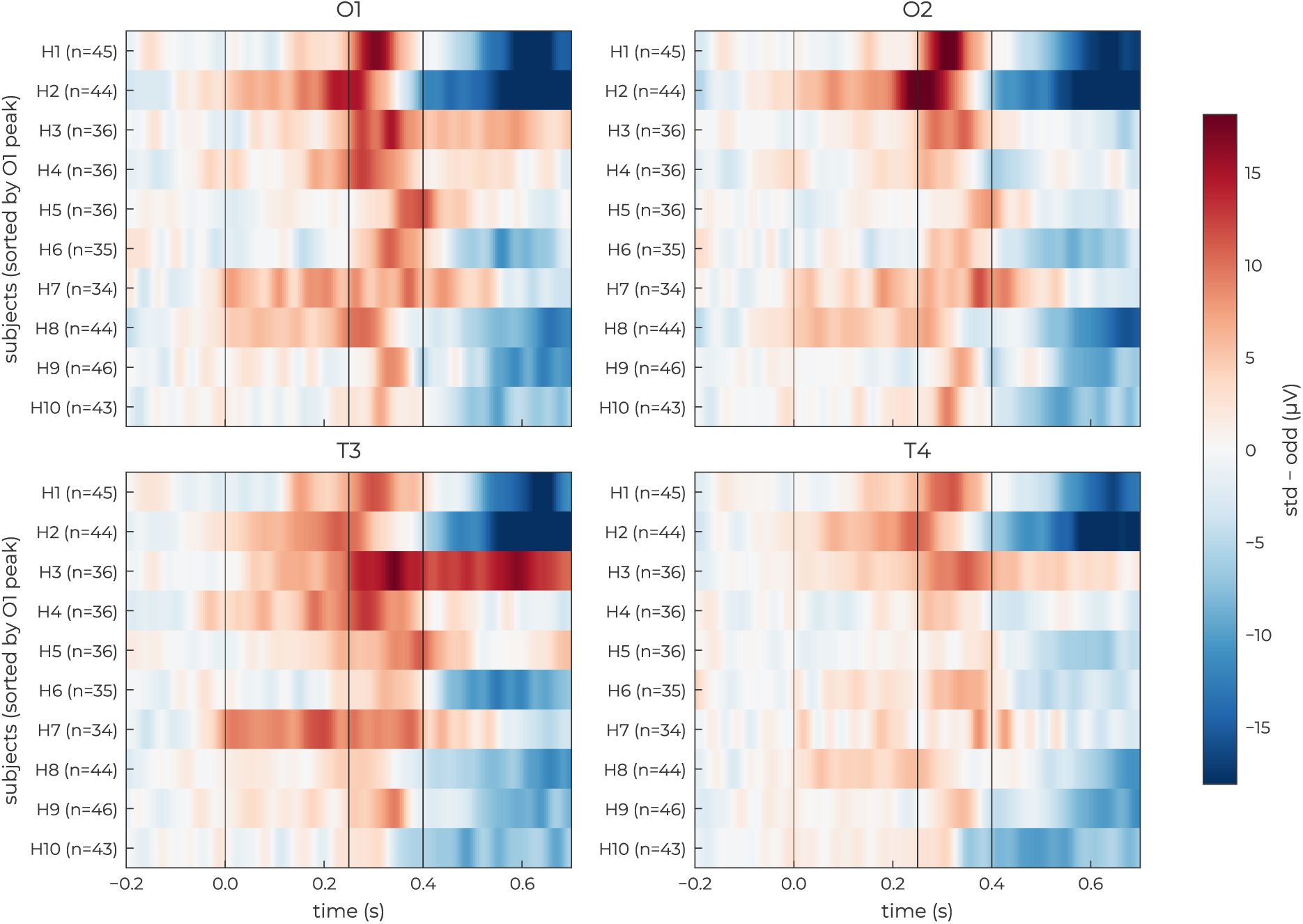
Per-subject difference waves. Rows: subjects sorted by O1 peak amplitude (strongest at top). Columns: time. Colour: amplitude in µV. Dotted vertical lines mark the 250–400 ms analysis window.

**Table 2:**
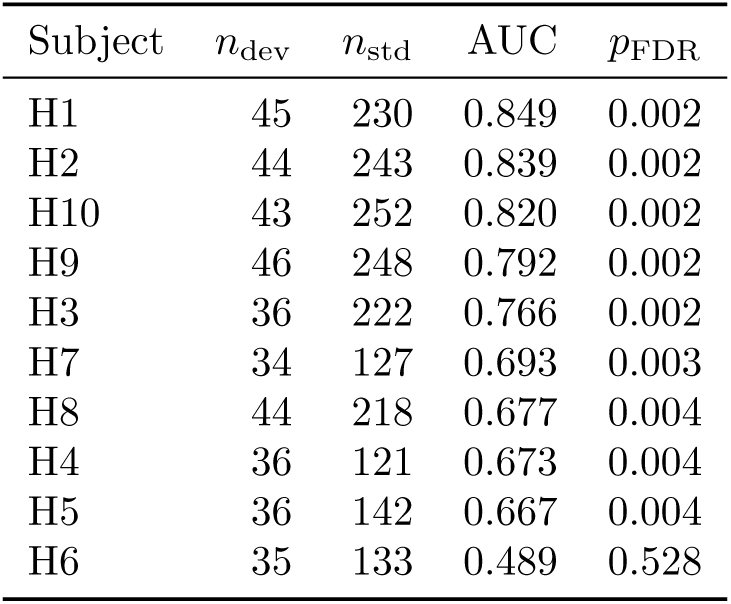
Per-subject cross-validated LDA AUC with permutation-based p-values (FDR-corrected across subjects). Subjects sorted by AUC.

At the individual level, a one-sided Welch’s *t*-test on epoch-level mean amplitude (deviant *<* standard, FDR-corrected across channels) detected a P300 on at least one channel in 8 of 10 subjects (95% Clopper–Pearson CI [44%, 97%]; exact binomial test against 5% chance, *p <* 10*^−^*^7^).

### 3.2 Trial-level classification

The LDA classifier separated deviant from standard trials above chance in 9 of 10 subjects (table 2; FDR-corrected). The group-level median AUC was 0.730 (bootstrap 95% CI [0.672, 0.820]; Wilcoxon vs. 0.5: p = 0.004). Per-subject AUC ranged from 0.489 (H6, the only subject not reaching significance) to 0.849 (H1); the distribution is shown in supplementary figure S3; per-subject ROC curves are in supplementary figure S4.

### 3.3 Clinical feasibility: MS patient

Patient P1 (relapsing–remitting MS) completed one game session on the headband hardware. The patient’s difference wave showed a clear positive deflection at all four channels (figure 6), with peak latencies inside the healthy cohort’s observed range (O1: 300 ms; O2: 300 ms; T3: 296 ms; T4: 324 ms). Peak amplitudes were at or above the healthy range (O1: 16.86 µV; O2: 15.30 µV; T3: 15.30 µV; T4: 14.49 µV); the T4 amplitude exceeded the healthy maximum (11.33 µV). Single-subject detection reached criterion on all four channels, and the trial-level LDA AUC of 0.853 exceeded the healthy group median of 0.730. Task accuracy was within the healthy range (green-gate collection rate 76.7%, cohort mean 84.7%, range [70.5, 97.2]%).

**Figure 6:**
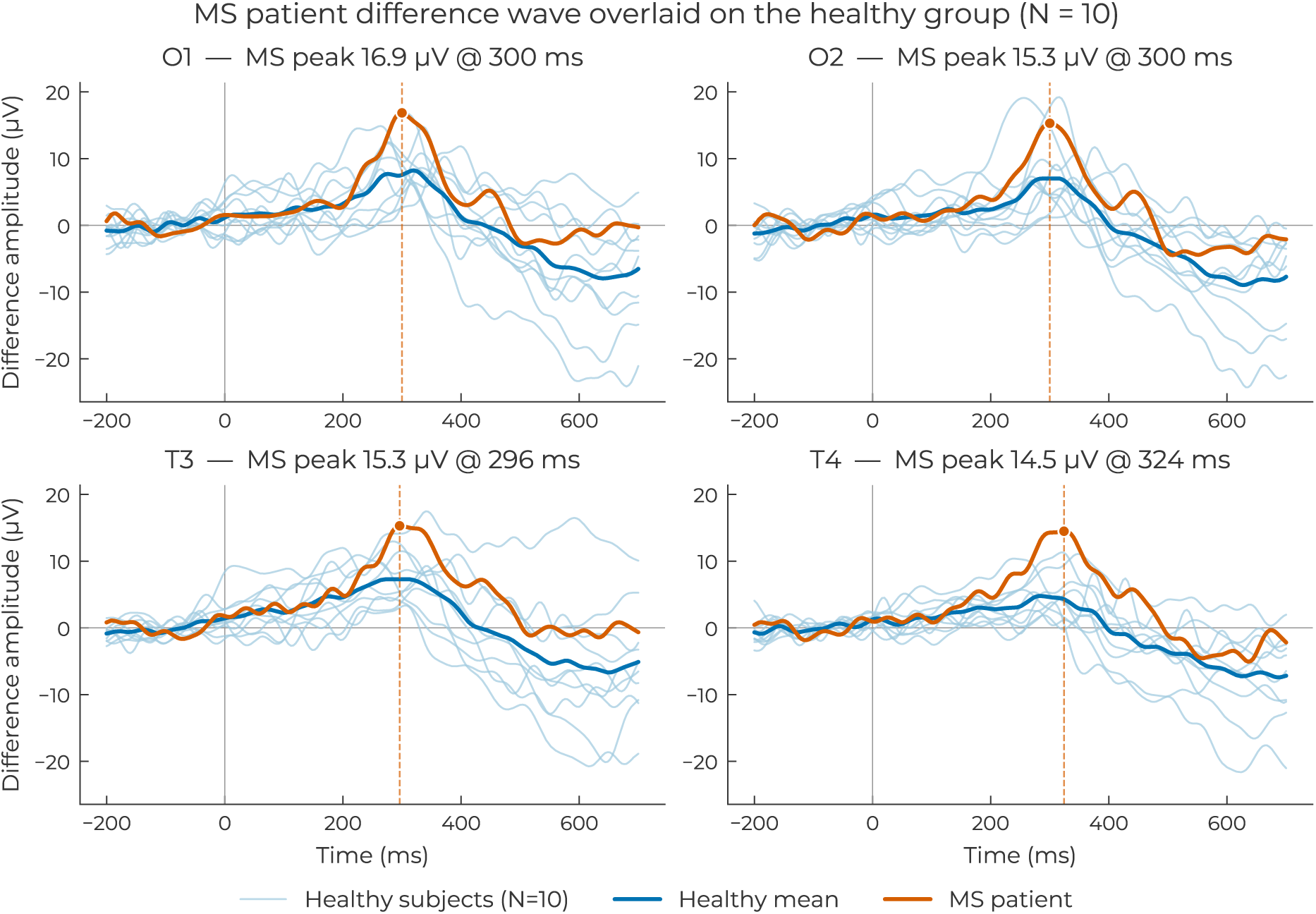
Difference wave of patient P1 (bold) overlaid on the healthy group grand-average (thin line with 95% CI band), per channel.

### 3.4 Game vs classical oddball

Subject H10 completed both the gamified task and six sessions of the classical visual oddball on the same headband hardware. The per-channel difference waves for the two paradigms are overlaid in figure 7 (the classical oddball grand-average is shown separately in supplementary figure S7). Waveform morphology was similar: the per-channel Pearson correlation of the two difference waves (across the full epoch) ranged from r = +0.61 (O2) to r = +0.76 (T4) (O1: +0.64; T3: +0.67). Per-channel peak amplitudes in the 250–400 ms window were larger in the game (O1: 6.86 µV, O2: 9.10 µV, T3: −5.76 µV, T4: −7.08 µV) than in the pooled classical sessions (O1: 3.19 µV, O2: −1.57 µV, T3: 1.29 µV, T4: −1.97 µV); peak latencies were comparable across paradigms and channels. Trial-level LDA AUC was 0.820 (p = 0.001) on the game session and 0.555 (p = 0.019) on the pooled classical sessions.

**Figure 7:**
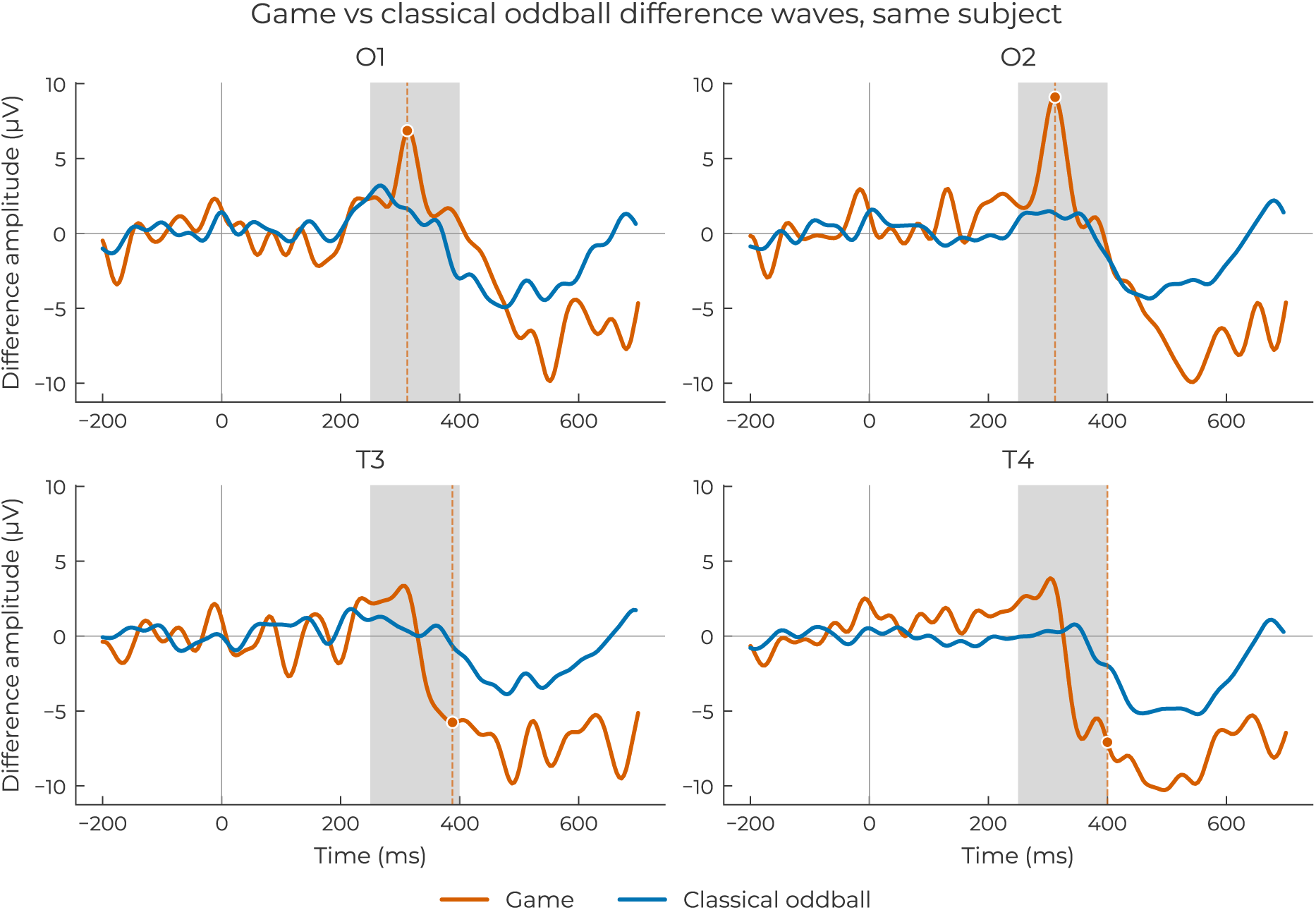
Per-channel difference-wave overlay for H10 on the gamified task (orange) and the classical visual oddball (blue). Yellow band: 250–400 ms analysis window.

### 3.5 Within-subject stability

Subject H10, the weakest P300 responder in the cohort, completed 48 game sessions. Single-session detection (one-sided Welch’s t-test, FDR-corrected) reached criterion in 5 of 48 sessions (95% CI [3%, 23%]). Trial-level classification was more sensitive: per-session LDA AUC exceeded chance in 34 of 48 sessions, with a median of 0.704. However, AUC showed a decreasing trend across session index (Spearman ρ = −0.71, p < 0.001; figure 8b).

**Figure 8:**
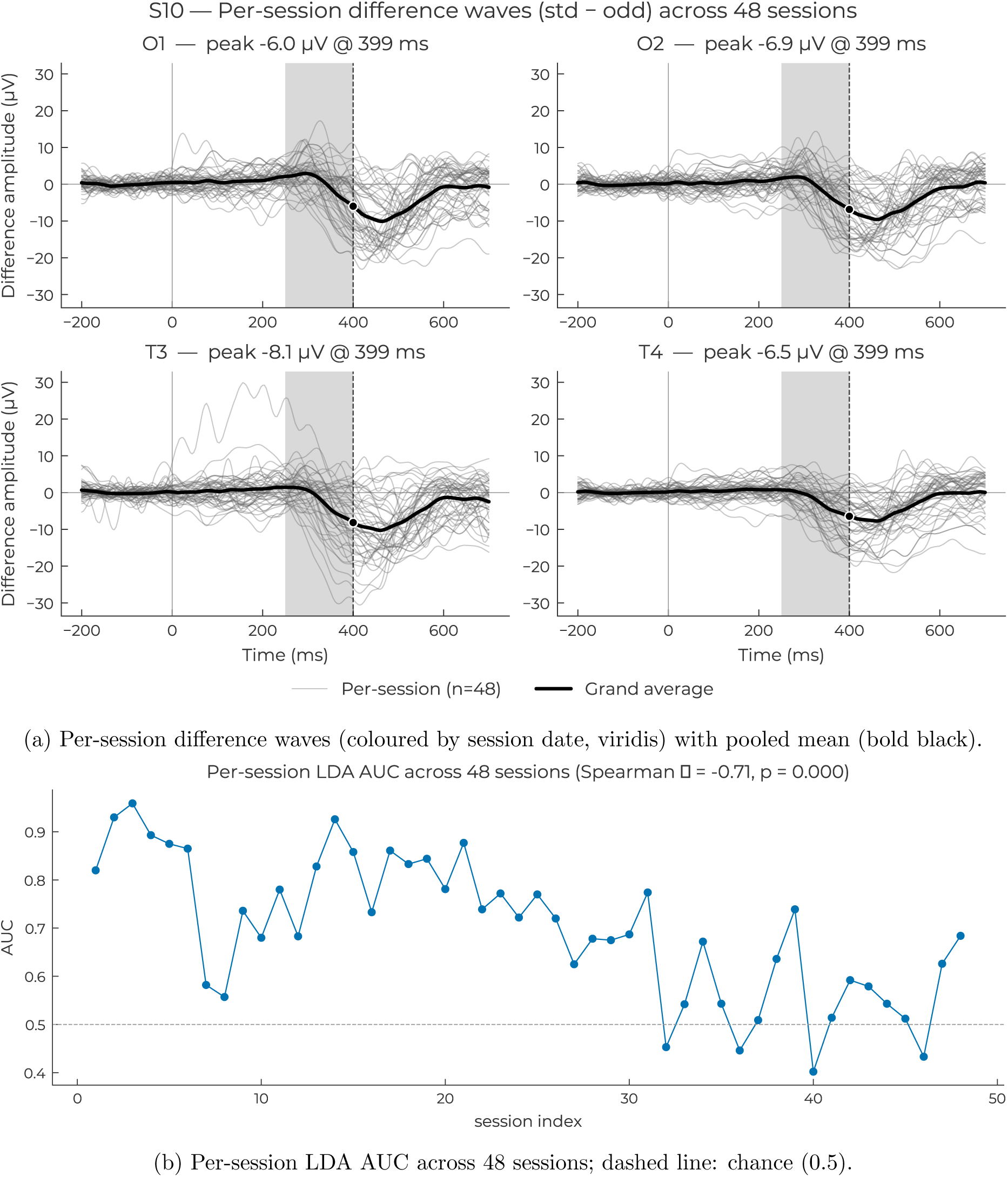
Within-subject stability for H10.

Within-session split-half reliability of the difference-wave peak was high on all channels (O1: 0.73; O2: 0.76; T3: 0.71; T4: 0.67). Between-session reliability, quantified as ICC(3,1) on per-session peak amplitudes, was near zero on all channels (O1: −0.05; O2: −0.11; T3: −0.25; T4: 0.06).

Subjects H1 (5 sessions) and H2 (3 sessions) showed qualitatively consistent waveform shapes across their additional sessions (supplementary figures S5 and S6).

### 3.6 Supplementary analyses

#### Headphone form factor

Two game sessions recorded with the EEG headphones (A1, A2, C3, C4) in subject H10 showed a P300-like deflection in the 300–400 ms window. Significant clusters (p < 0.05, trial-level permutation test) emerged on channels A2 and C4 in one of the two sessions. Per-session ERPs are shown in supplementary figure S8, and a direct overlay of headphones and headband difference waves from the same subject is in supplementary figure S9.

#### Behavioural–neural correlation

No correlation between task accuracy and difference-wave peak amplitude reached significance after FDR correction (0 of 12 tests across channels and behavioural measures). Correlations between LDA AUC and task performance were similarly weak, with all bootstrap 95% CIs including zero. The narrow range of task accuracy in this cohort (all subjects collected green gates at high rates; supplementary figures S10 and S11) limits the power to detect such relationships.

## 4 Discussion

The central finding of this study is that P300 can be recorded as a gameplay-integrated neural response during genuine gameplay using a four-channel wearable EEG headband. As outlined in the Introduction, prior P300 gaming paradigms use the component as a voluntary BCI control signal. The present study takes a different approach: the oddball stimuli were integral to the game mechanics, and subjects played the game while the P300 emerged from the natural cognitive demands of the task itself. Consequently, the P300 observed here reflects natural cognitive processing during an engaging task, not effortful compliance with experimental instructions, nor with a BCI paradigm in which P300 serves as the control signal.

### 4.1 Signal quality and the role of engagement

The per-subject peak amplitudes observed here (O1: 11.76 µV, O2: 10.47 µV, T3: 8.73 µV, T4: 6.09 µV) exceeded what has typically been reported with comparable consumer-grade hardware. Debener et al. (2015) demonstrated reliable P300 detection using around-the-ear cEEGrid electrodes with a standard oddball, and Lee et al. (2024) found that consumer wireless EEG devices produced detectable but temporally distorted P300 waveforms. Two factors likely contribute to the comparatively large amplitudes in the present study. First, the forehead reference, positioned on the opposite side of the head from the recording electrodes, may capture a larger potential difference than standard mastoid or earlobe references. This would be a geometrical consequence of reference placement (consistent with the re-referencing effects documented by Jovanović et al. 2025 for the N400), not a signal quality advantage per se, but it increases the measured amplitude and may improve signal-to-noise ratio for downstream classification. Second, the gamified context itself may sustain greater attentional engagement than repetitive oddball tasks. Pei et al. (2025) identified a P300-like component during gameplay characterised by unusually large magnitude and no habituation across repeated exposures, suggesting that well-designed game mechanics may counteract the neural adaptation that typically attenuates P300 with stimulus repetition. The large amplitudes observed here are consistent with both mechanisms operating simultaneously.

The observation that occipital channels showed larger effects than temporal channels aligns with recent evidence that parieto-occipital electrodes are among the strongest contributors to P300 classification (Kim and Kim, 2025). While the absence of parietal coverage (Pz, Cz) remains a limitation, the headband’s occipital placement captures the posterior tail of the P300 topography, as confirmed by the group-level detection on O1 and O2 and the classification results.

The classification performance (group median AUC 0.730, 9 of 10 subjects above chance) should be interpreted in the context of the hardware constraints. Ganin et al. (2013) reported 65% single-trial accuracy with a 6-channel posterior montage and moving stimuli in a P300 BCI, and Guger et al. (2012) achieved above 90% accuracy with an 8-electrode intendiX P300 speller. That comparable discrimination was achieved here with four channels at non-standard positions, during a task not designed for classification, suggests the gamified P300 carries sufficient discriminative information for single-trial detection despite the hardware constraints. The paradigm was not intended for real-time command selection, and its information transfer rate would be insufficient for BCI control. For passive monitoring applications requiring binary detection (P300 present vs. absent), an AUC of 0.730 provides sufficient separation to identify whether a P300 was elicited in a given session, a separation sufficient for session-level detection in passive monitoring applications.

### 4.2 Convergent evidence across paradigms and form factors

Two supplementary comparisons reported in the Results, each for a single subject, deserve interpretation because they address the generalisability of the main findings from different angles. The within-subject comparison between the gamified and classical visual oddball (H10) revealed highly similar waveform morphology, suggesting that the gamified paradigm may engage the same neural generator as the standard oddball. The game environment introduces continuous visual motion, dynamic backgrounds, and attentional competition from multiple on-screen elements, all of which could in principle alter or suppress the P300 response. The preserved waveform shape indicates that the oddball constraints embedded in the game design (deviant-to-standard ratio, stimulus timing, pseudo-randomised spatial positions) appear to have been sufficient to maintain the psychophysiological conditions for P300 elicitation in this subject despite the increased visual complexity.

The direction of the classification difference is informative: trial-level LDA AUC was 0.820 on the game session compared to 0.555 on the pooled classical oddball sessions, despite the classical dataset containing far more trials. With a single subject, this observation cannot be generalised, but it is consistent with the hypothesis that gamification enhances rather than degrades P300 discriminability. The classical oddball is, by design, monotonous, and the findings of Pei et al. (2025) would predict this pattern: a game that sustains attention may produce a more discriminable P300 than a stripped-down laboratory task. Cattan et al. (2020) identified low information transfer rate and stimulation-induced fatigue as fundamental barriers for P300 BCI in gaming environments; the present paradigm sidesteps these problems because P300 is not the control signal, and collecting the target stimuli (green gates) is itself the core game mechanic, making the task intrinsically engaging.

The two pilot sessions with the headphone form factor showed a P300-like deflection, with significant clusters on two channels in one session. Though preliminary, this result suggests that the gamified P300 is detectable with a central-channel montage. The signal can be recorded from electrode positions near the ears and over central cortex. This form-factor versatility has practical implications: if P300 monitoring can be performed with both headband and headphones, the barrier to longitudinal data collection is lowered by offering users a choice of wearable configurations.

### 4.3 Polarity inversion: reference geometry, not artifact

The headband uses a forehead reference, which differs from the standard mastoid or earlobe configurations used in the P300 literature. Although the P300 is maximal at parietal sites, its broad scalp distribution extends frontally (Polich, 2007): the P300 voltage at the forehead exceeds that at occipital and temporal sites. Consequently, when occipital or temporal electrodes are referenced to the forehead, the recorded signal (V_electrode_ − V_forehead_) is negative during the P300 window, inverting the polarity relative to what is observed with standard references. In the present data, the difference wave computed as deviant minus standard produced a negative deflection in the P300 window. To restore the conventional positive morphology, the difference wave throughout this paper is computed as standard minus deviant. This convention was adopted precisely because it yields waveforms that facilitate comparison with the existing literature.

Three converging lines of evidence support the interpretation that the observed polarity inversion is a deterministic consequence of reference geometry rather than an artifact of the gamified paradigm. First, the same inverted polarity was observed in the classical visual oddball recorded with the same headband in the same subject, confirming that the inversion is not specific to the game environment. Second, the headphone form factor, which uses a different reference location (at the top centre of the headband, approximately at Cz, rather than at the forehead), produced the expected positive P300 in the deviant-minus-standard difference wave. The fact that moving the reference away from the forehead restores the conventional polarity constitutes a direct within-device-family validation that the effect is driven by reference location. Third, the polarity inversion is consistent with the findings of Jovanović et al. (2025), who demonstrated that re-referencing can substantially alter ERP morphology, including polarity reversals, in the case of the N400 component. Across all three configurations (gamified headband, classical headband, and gamified headphones) the deviant–standard difference is concentrated in the same 250–400 ms window, with only the polarity differing between forehead-referenced and Cz-referenced recordings; the temporal signature is therefore consistent with a P300 component, with the apparent sign of the wave determined by the reference location rather than by the underlying neural process.

The implication is general: any wearable EEG device that uses a forehead reference will produce P300 waveforms with inverted polarity at posterior and temporal sites. Researchers and clinicians using such devices should anticipate this inversion and account for it in their analysis pipelines, rather than interpreting it as evidence against P300 elicitation.

### 4.4 Clinical feasibility

The MS patient produced a clear P300 with amplitudes at the upper end of the healthy range and classification performance exceeding the healthy group median. This direction of effect is unexpected given the clinical literature, where P300 amplitude is typically reduced and latency prolonged in MS (Kaddoori, 2023). Several considerations apply. The patient had relapsing-remitting MS, the subtype most likely to retain cognitive function, and P300 amplitude in RRMS has been shown to relate to preserved cognition rather than disease severity (Sundgren et al., 2015). With a single patient and a single session, no clinical conclusions can be drawn. The result establishes only that the paradigm is tolerable and produces a measurable signal in this population, which is the minimal requirement for proceeding to a powered clinical study.

Given the established sensitivity of P300 to cognitive status in MS and the advantage of visual over auditory paradigms (see Introduction), a wearable approach enabling unsupervised P300 elicitation during short game sessions at home could transform cognitive monitoring from episodic clinical visits into continuous data collection. Whether the present paradigm is sensitive to the specific P300 changes documented in MS requires investigation with a larger, more heterogeneous patient cohort.

### 4.5 Longitudinal stability and the limits of repeated measurement

The 48-session dataset from H10 provides unusually dense longitudinal P300 data. Most P300 studies report one or two sessions per subject, and longitudinal data of this density, recorded under unsupervised home conditions during genuine gameplay, are essentially absent from the literature.

The pattern of results is mixed. Within-session reliability was high (split-half correlations exceeding 0.70 on three of four channels), meaning the signal is internally consistent within a single game session. Between-session reliability, by contrast, was essentially zero (ICC near 0 on all channels). The P300 amplitude on any given session was not predictive of the amplitude on the next.

This dissociation is not unique to the gamified paradigm. P300 amplitude fluctuates with arousal, attention, time of day, and recent cognitive load, all of which vary between sessions in an unsupervised home setting. The near-zero ICC may be better understood as a property of P300 variability under ecological conditions than as a failure of the paradigm. For monitoring applications, the practical consequence is that a single session should not be treated as a reliable individual measurement. Estimates should be pooled across multiple sessions, with the number calibrated to the desired precision.

The declining AUC trend across H10’s 48 sessions has different implications depending on the intended application. If P300 is used as a cognitive biomarker (e.g., tracking disease progression in MS), the paradigm must remain constant across sessions so that any change in P300 parameters can be attributed to the patient’s cognitive state, not to the task. In this case, the declining trend is a confound that needs to be characterised and controlled for, and changing the game content between sessions would introduce an additional source of variance that undermines the biomarker interpretation. If, on the other hand, P300 discriminability is treated as an engagement marker, the declining trend is itself informative, and the factors driving it (habituation, motivational decline, electrode fit, time of day) become the variables of interest. These two use cases impose opposing design constraints: biomarker stability requires a fixed paradigm, while engagement monitoring benefits from understanding and potentially manipulating the sources of session-to-session variation.

The contrast with H3 and H5, who showed qualitatively consistent waveforms across their additional sessions, suggests that stability is better in stronger P300 responders. A larger signal-to-noise ratio produces more stable per-session estimates, as expected. For the monitoring use case, most users (those with clearly detectable P300) may require fewer sessions for stable estimates than H10, the weakest responder in the cohort.

### 4.6 Early component suppression

The grand-average waveforms showed attenuated early visual components (N1/P2 complex, first 200 ms) relative to what is typically reported in classical oddball studies with research-grade EEG. The within-subject comparison in H10 provides a direct test of whether this attenuation is driven by the game environment or by the hardware configuration. In the present study, the classical oddball also showed attenuated early components, suggesting that the suppression is attributable to the forehead reference and limited hardware bandwidth rather than to the visual complexity of the game environment. This interpretation is consistent with the general observation that early sensory components are more susceptible to recording conditions than later cognitive ERPs (Scanlon et al., 2019). In either case, the P300 window (250–400 ms) was preserved across both paradigms, indicating that the later cognitive component is more resistant to whichever factor suppresses the early sensory response.

### 4.7 Design principles for gamified ERP paradigms

Three design principles were central to preserving P300 elicitation within engaging gameplay, and they generalise beyond this specific implementation.

First, the deviant and standard probabilities were fixed at approximately 0.2 and 0.8 (empirical 0.198/0.802 across the cohort), consistent with standard oddball paradigms, and the stimulus timing preserved an interval sufficient for P300 recovery between successive deviants. These constraints are non-negotiable for any oddball-based paradigm, gamified or not.

Second, the temporal separation and laterality randomization between stimulus onset and motor response were exploited to decouple cognitive and motor activity. Because the avatar’s running speed was fixed, the P300 response window (250–400 ms after gate colour reveal) preceded the motor response (lane change to collect or avoid the gate). Gate positions were pseudo-randomised across lanes, ensuring that any residual movement-related potentials were distributed equally across stimulus categories and cancelled during ERP averaging.

Third, the game mechanics introduced variety (environments, obstacles, scoring) to counteract the monotony inherent in oddball paradigms. The declining AUC trend in H10’s extended recordings suggests that even gamification may have limits in sustaining engagement, though the design implications depend on the intended application (see Section 4.5).

These principles (ratio preservation, motor-confound decoupling, engagement maintenance) define the design space for any attempt to embed ERP paradigms into interactive applications. The present study provides empirical evidence that this balance is achievable.

### 4.8 Limitations and future directions

The sample is small (N=10) and demographically narrow (eight male, two female; aged 28–37; all right-handed), and individual variability was substantial (AUC range 0.489–0.849). The absence of parietal coverage — where P300 amplitude is maximal (Polich, 2007) — likely explains the two participants in whom individual-level detection failed; wearable devices with parietal channels should improve detection rates, and the headphone pilot provides initial evidence that non-headband form factors are viable. The game-versus-classical comparison and the headphones recordings each rest on a single participant and must not be generalised. The MS patient provides feasibility evidence only; clinical utility requires a powered clinical cohort. The behavioural–neural correlation analysis yielded null results throughout, but a ceiling effect in task accuracy — nearly all participants collected green gates at high rates — precluded any meaningful test; difficulty titration in future designs would allow this question to be revisited. Finally, whether the suppression of early visual components (N1/P2) reflects the forehead reference, the hardware bandwidth, or the visual complexity of the game remains unresolved, because the classical oddball showed the same suppression with the same device; a direct comparison with a research-grade system at matched electrode positions would isolate these contributions. Beyond a larger replication study, several extensions are motivated directly by the present findings. The gamification principles validated here are not specific to P300: the mismatch negativity (MMN), elicited by unattended deviants without any overt response, may be embedded even more seamlessly into gameplay and is an attractive target for unsupervised monitoring applications. A within-subject design comparing fixed and periodically refreshed game versions could distinguish neural habituation from motivational decline in the longitudinal AUC trend. And a powered MS study, with repeated assessments across the relapsing–remitting cycle, would test whether the paradigm is sensitive to the specific cognitive fluctuations for which P300 has established biomarker value (Kaddoori, 2023; Vlieger et al., 2024) — the clinical question that motivated this work.

## 5 Conclusion

A wearable, four-channel EEG headband, self-mounted at home, captures a robust P300 during genuine gameplay. The signal is detectable at the individual level in the majority of participants, classifiable above chance on single trials, morphologically consistent with classical oddball responses, and present in a patient with multiple sclerosis. Gamification does not compromise P300 elicitation; in the within-subject comparison reported here, it improved it. Between-session variability under unsupervised ecological conditions demands multi-session averaging for reliable individual measurement. These findings establish the empirical foundation and identify the practical constraints for ambulatory, gamified P300 monitoring — a scalable approach to cognitive assessment that requires no laboratory, no technician, and no break from everyday life.

## Data Availability

The data generated in this study are not publicly available.

## Author Contributions

**Conceptualization:** A.M.S., B.S. and Z.T.; **Methodology:** A.M.S., B.S.; **Software:** S.G., Z.T., A.M.S.; **Formal analysis:** B.S., A.M.S.; **Investigation:** B.S., A.M.S.; **Resources:** Z.T.; **Data curation:** B.S.; **Writing (original draft):** B.S., A.M.S.; **Writing (review & editing):** all authors; **Visualization:** B.S.; **Supervision:** D.K., R.S., A.M.S., Z.T.; **Project administration:** Z.T., S.G.; **Funding acquisition:** Z.T.

## Acknowledgment

The authors would like to thank the Ministry of the Economy in Luxembourg and its digital health directorate for co-supporting this research. Similarly, we would like to thank the Luxembourg National Research Fund (FNR) for partially funding this research.

## Supplementary Materials

**Figure S1:**
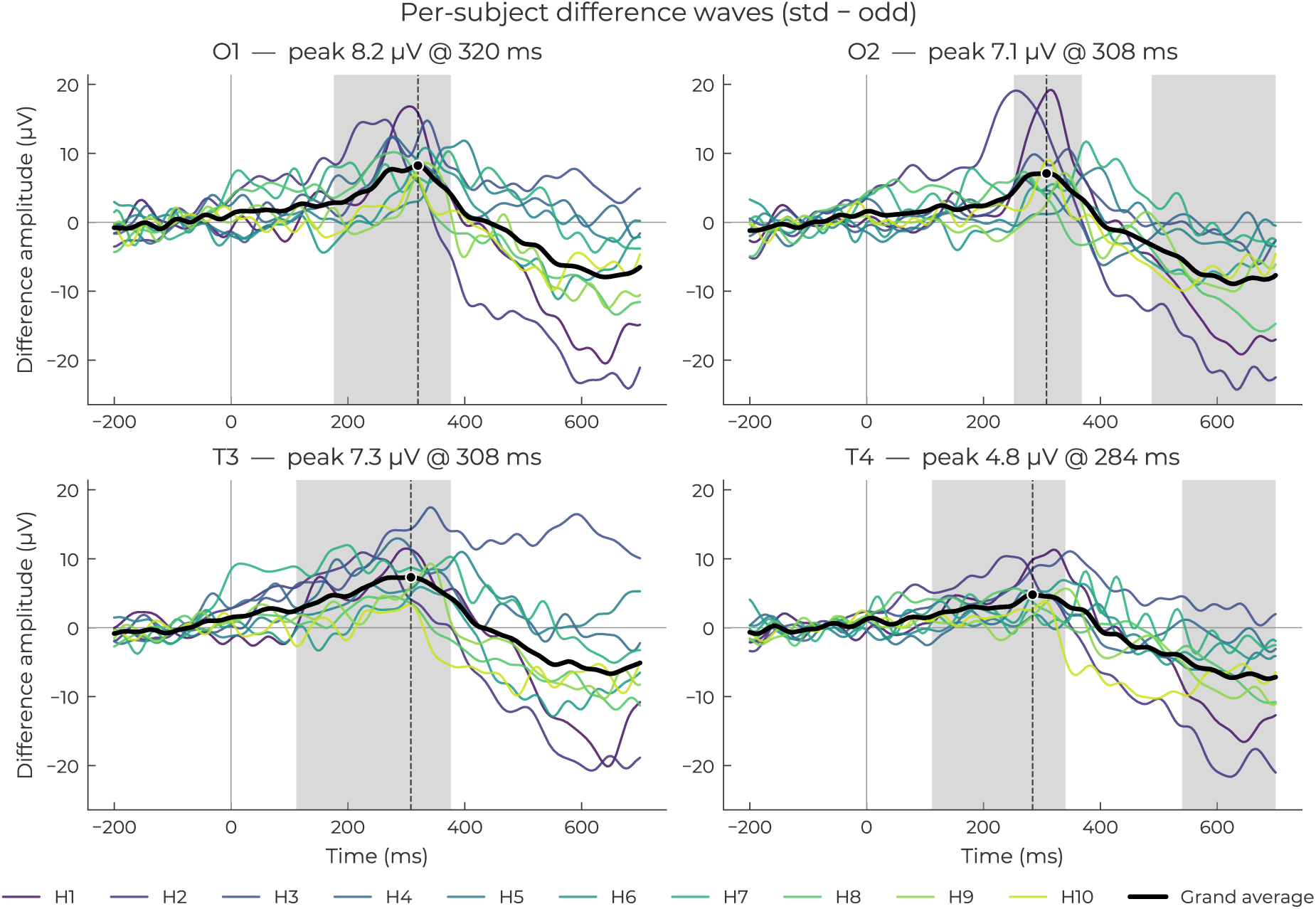
Per-subject difference waves (standard minus deviant) for all 10 healthy subjects (thin lines) with grand-average overlay (bold black). Shaded region: P300 window (250–400 ms). Each panel corresponds to one channel (O1, O2, T3, T4).

**Figure S2:**
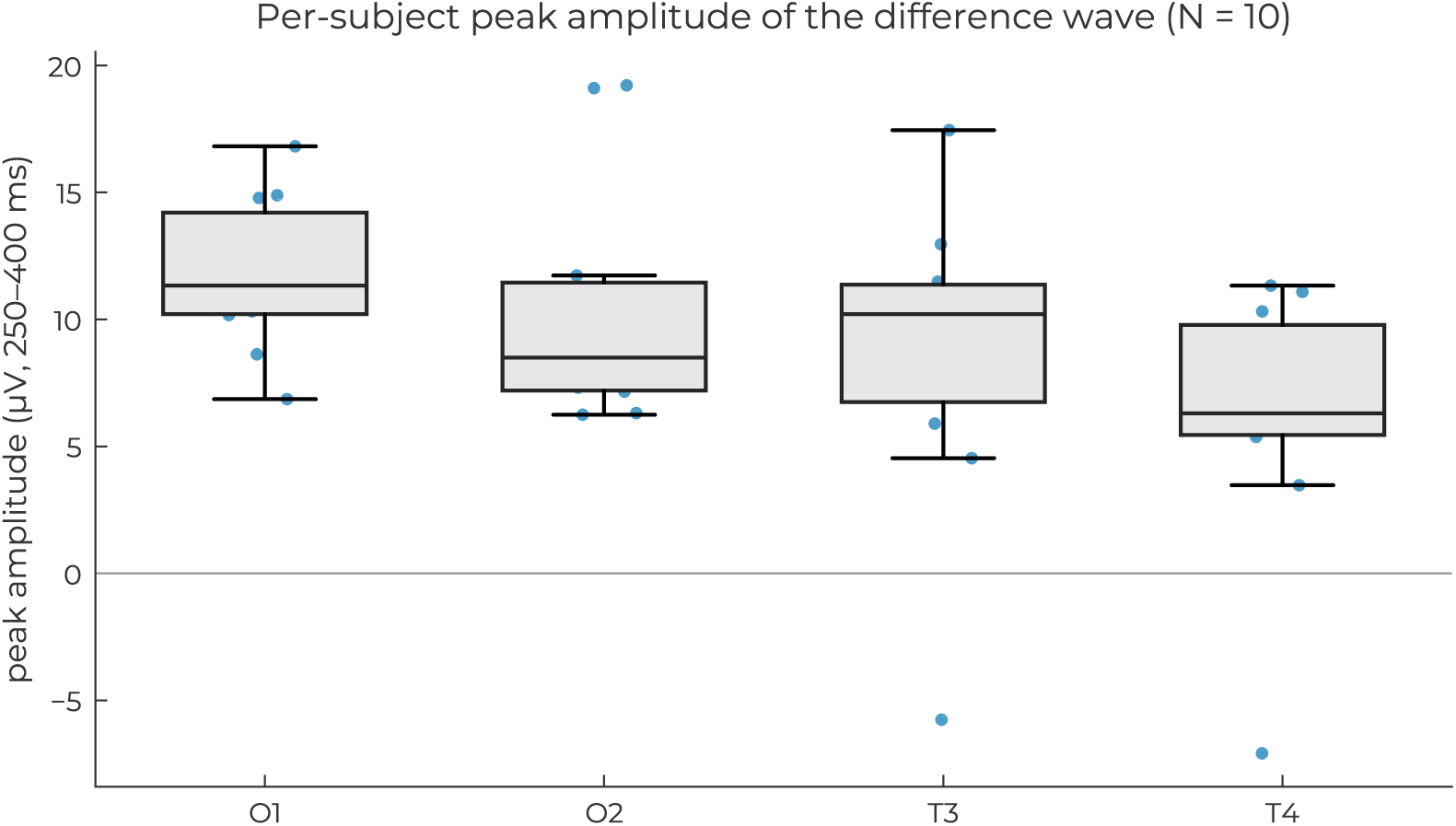
Per-subject difference-wave peak amplitude (left) and latency (right) in 250–400 ms, per channel. Coloured dots: individual subjects; box: interquartile range; whiskers: 1.5×IQR; black line: median.

**Figure S3:**
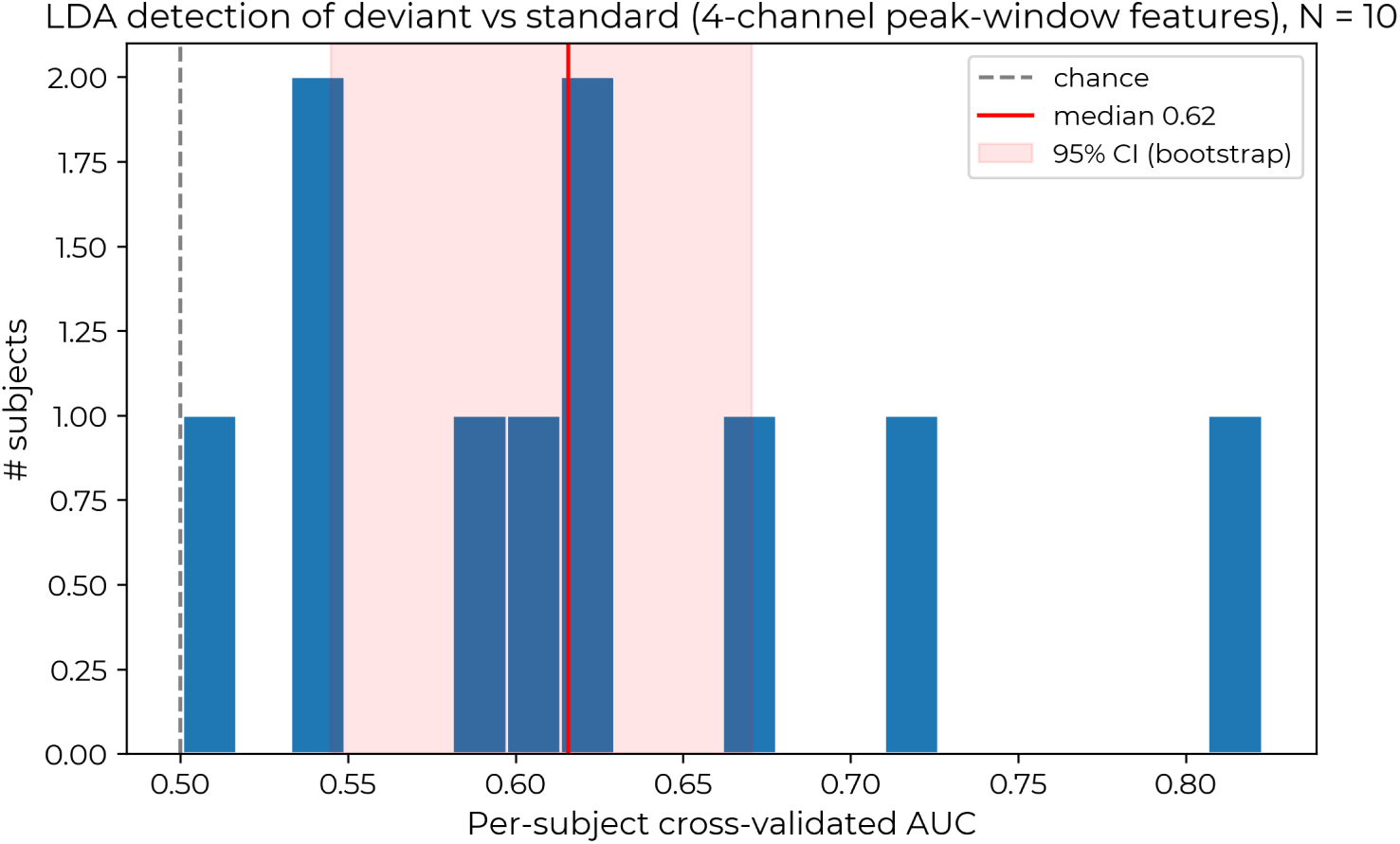
Distribution of per-subject cross-validated LDA AUC values (N = 10). Dashed black line: chance level (0.5). Red line: group median. Shaded band: 95% bootstrap confidence interval.

**Figure S4:**
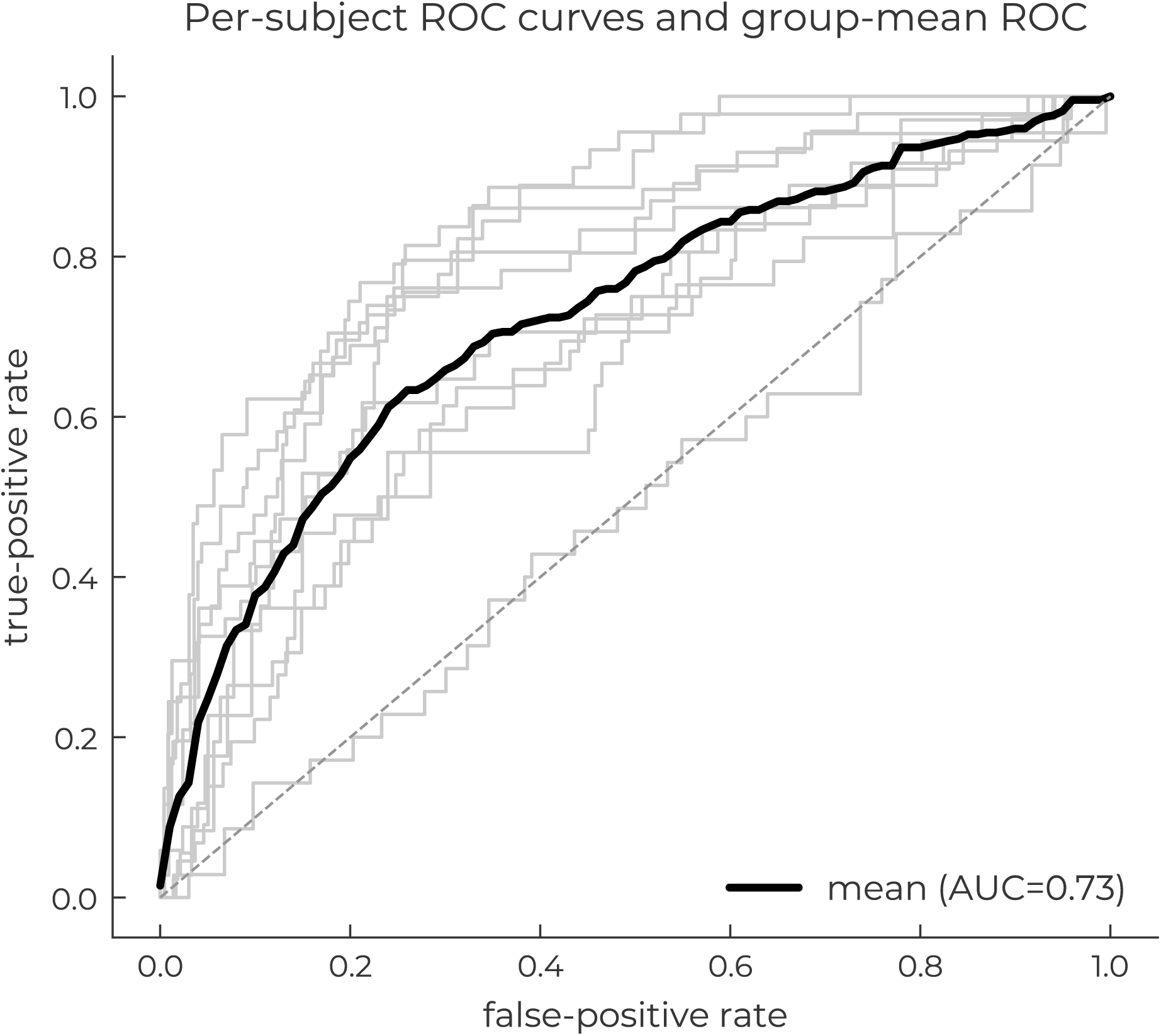
Per-subject cross-validated ROC curves (grey) with the across-subject mean ROC (bold black). Red dashed: chance.

**Figure S5:**
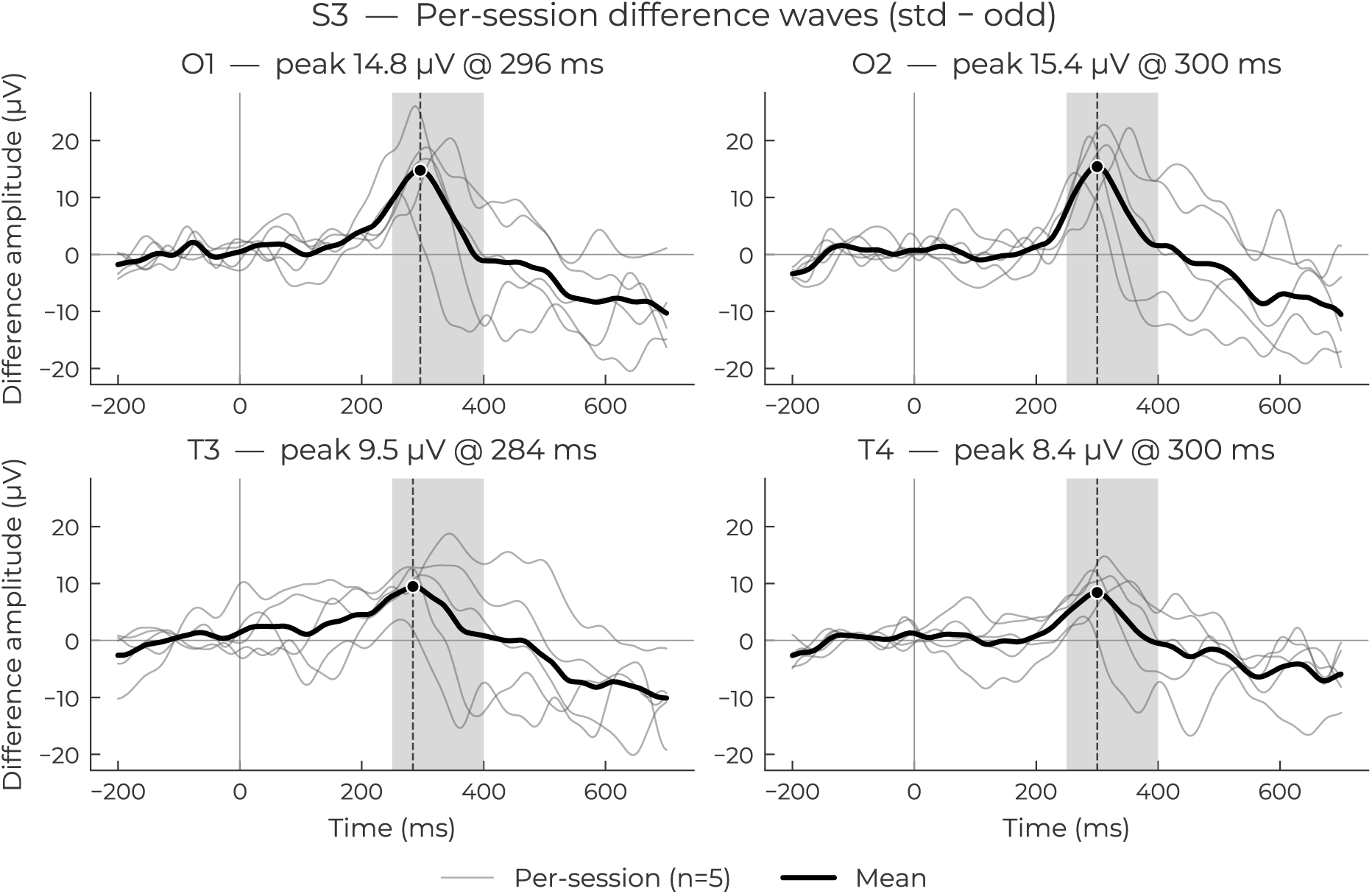
Within-subject stability for H1 (5 sessions). Per-session difference waves with pooled mean (bold black). H1 was the strongest P300 responder in the cohort, showing qualitatively more reproducible peak amplitudes and latencies than H10.

**Figure S6:**
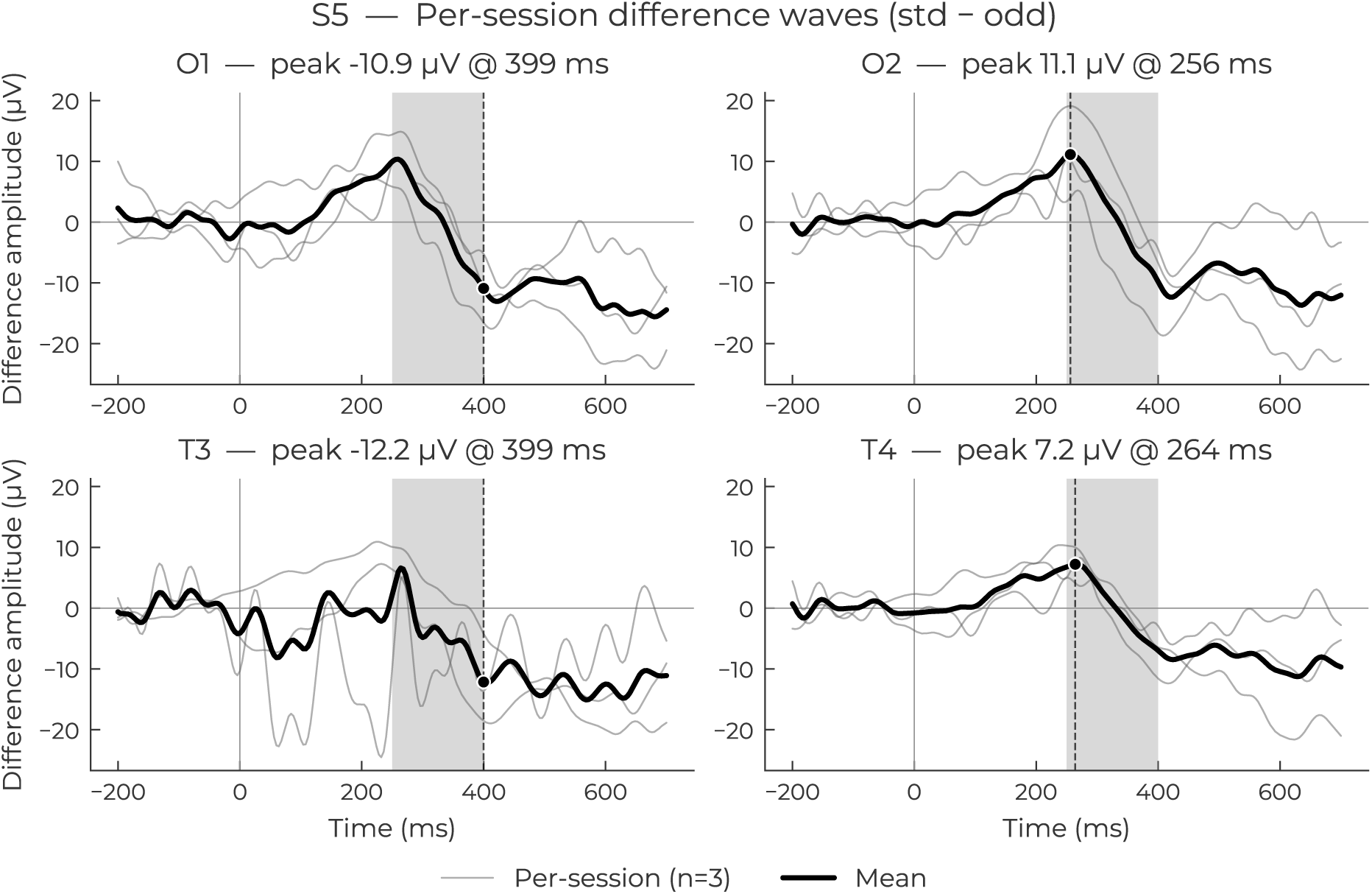
Within-subject stability for H2 (3 sessions). Per-session difference waves with pooled mean (bold black).

**Figure S7:**
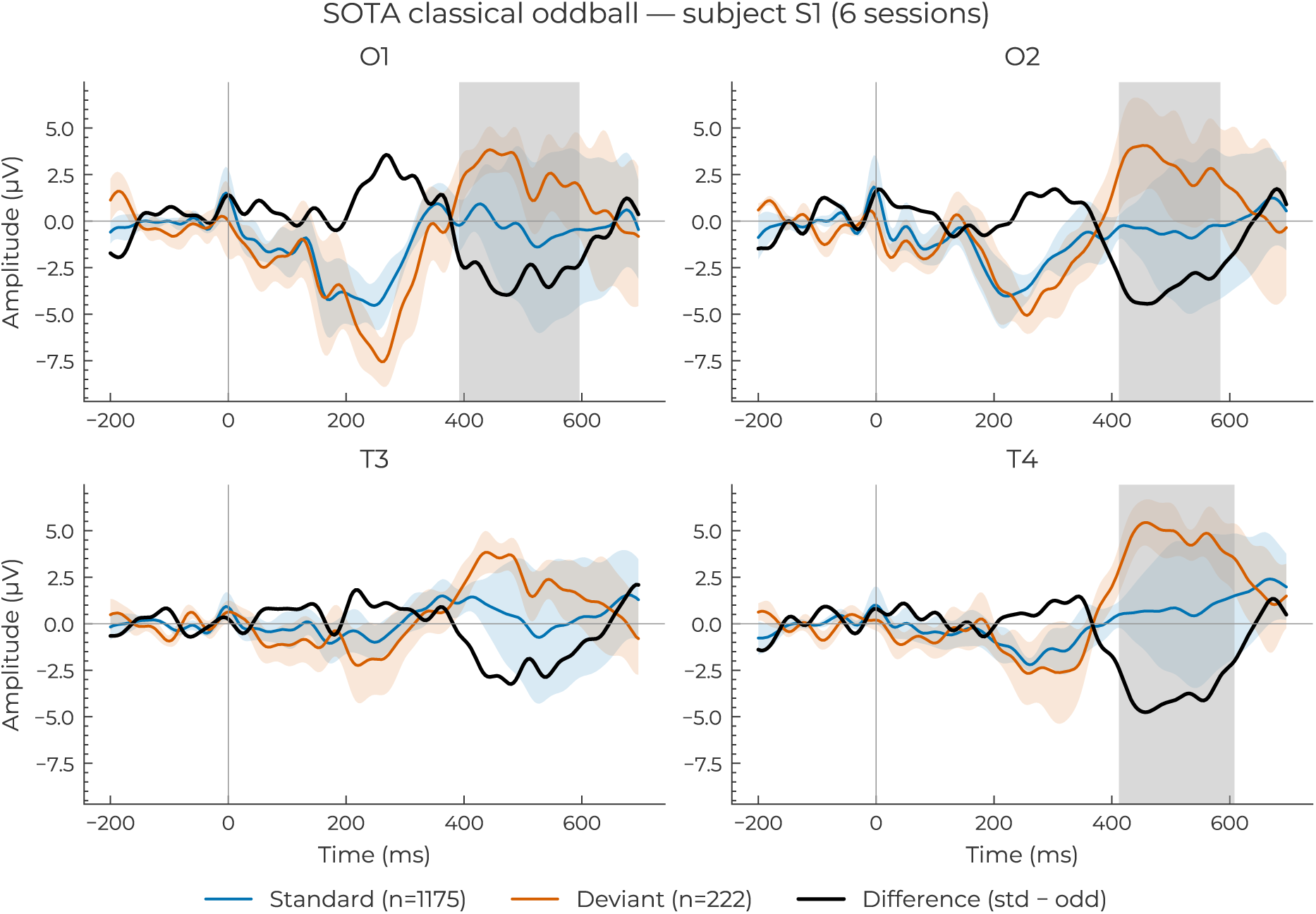
Grand-average ERP for the classical visual oddball (6 sessions, subject H10). Blue: standard; red: deviant; black: difference wave (standard minus deviant).

**Figure S8:**
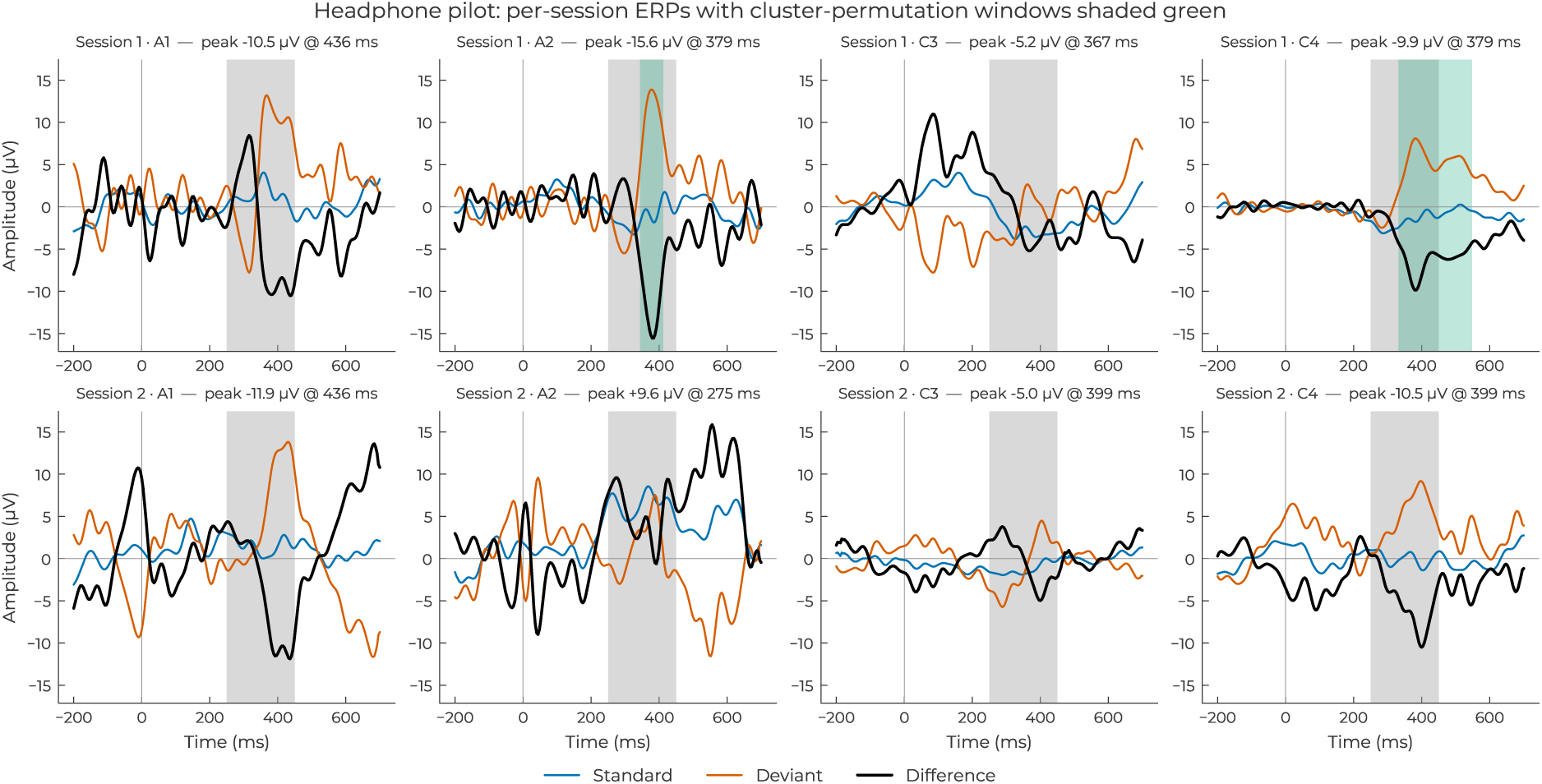
Headphone form factor: per-session per-channel ERPs (standard, deviant, difference). Grey bands mark significant clusters from a trial-level two-sample permutation test (p < 0.05, 2000 permutations).

**Figure S9:**
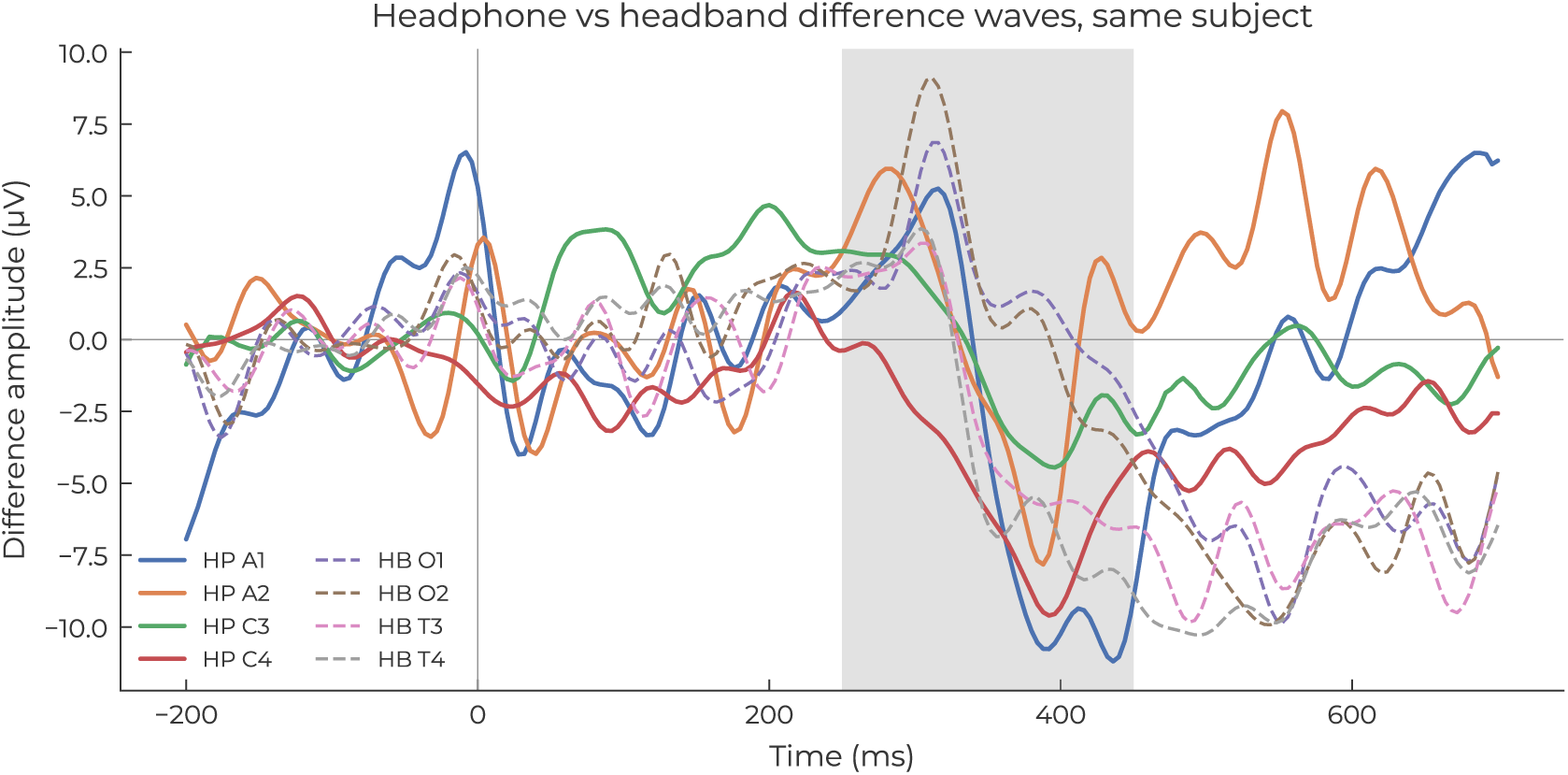
Same subject’s difference wave recorded with the headphones (solid, A1/A2/C3/C4) and headband (dashed, O1/O2/T3/T4) montages, paired by laterality for side-by-side comparison. Electrodes are anatomically distinct; the pairings are visual aids, not equivalence claims.

**Figure S10:**
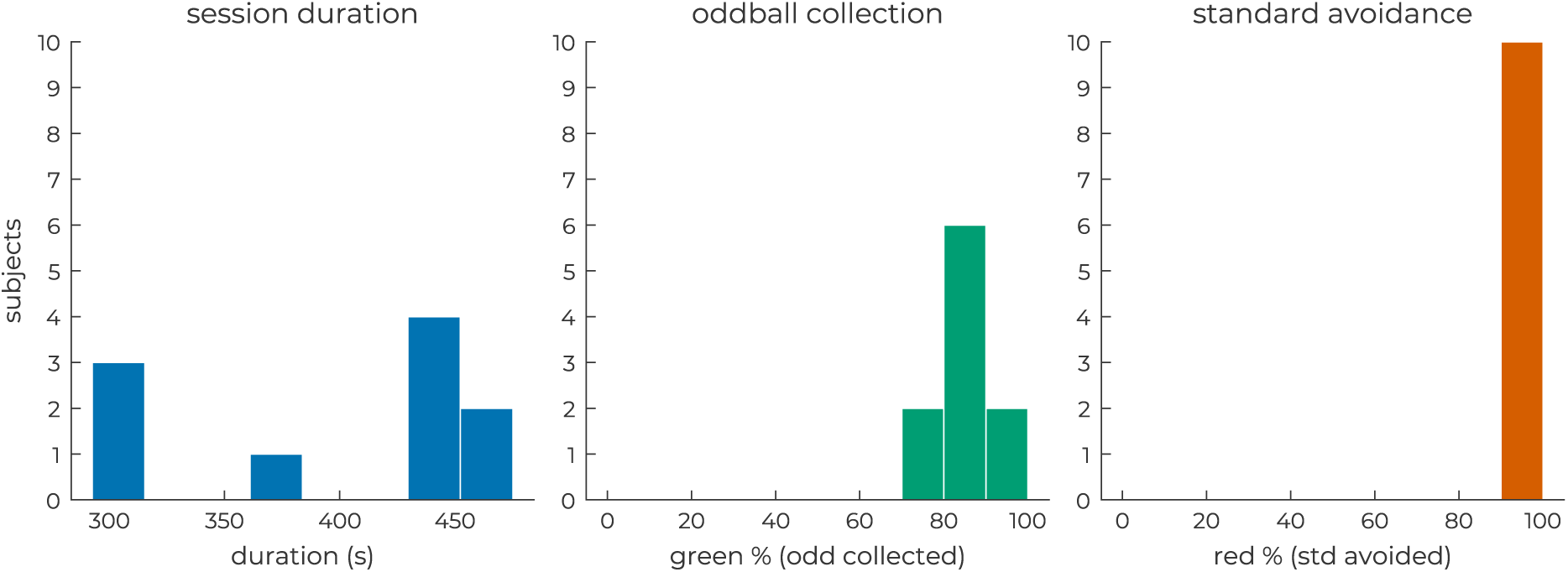
Distribution of per-session oddball task accuracy (green-gate collection rate) across all sessions. Dashed lines indicate threshold levels. The majority of sessions showed high col-lection rates, consistent with the ceiling effect discussed in the behavioural–neural correlation analysis.

**Figure S11:**
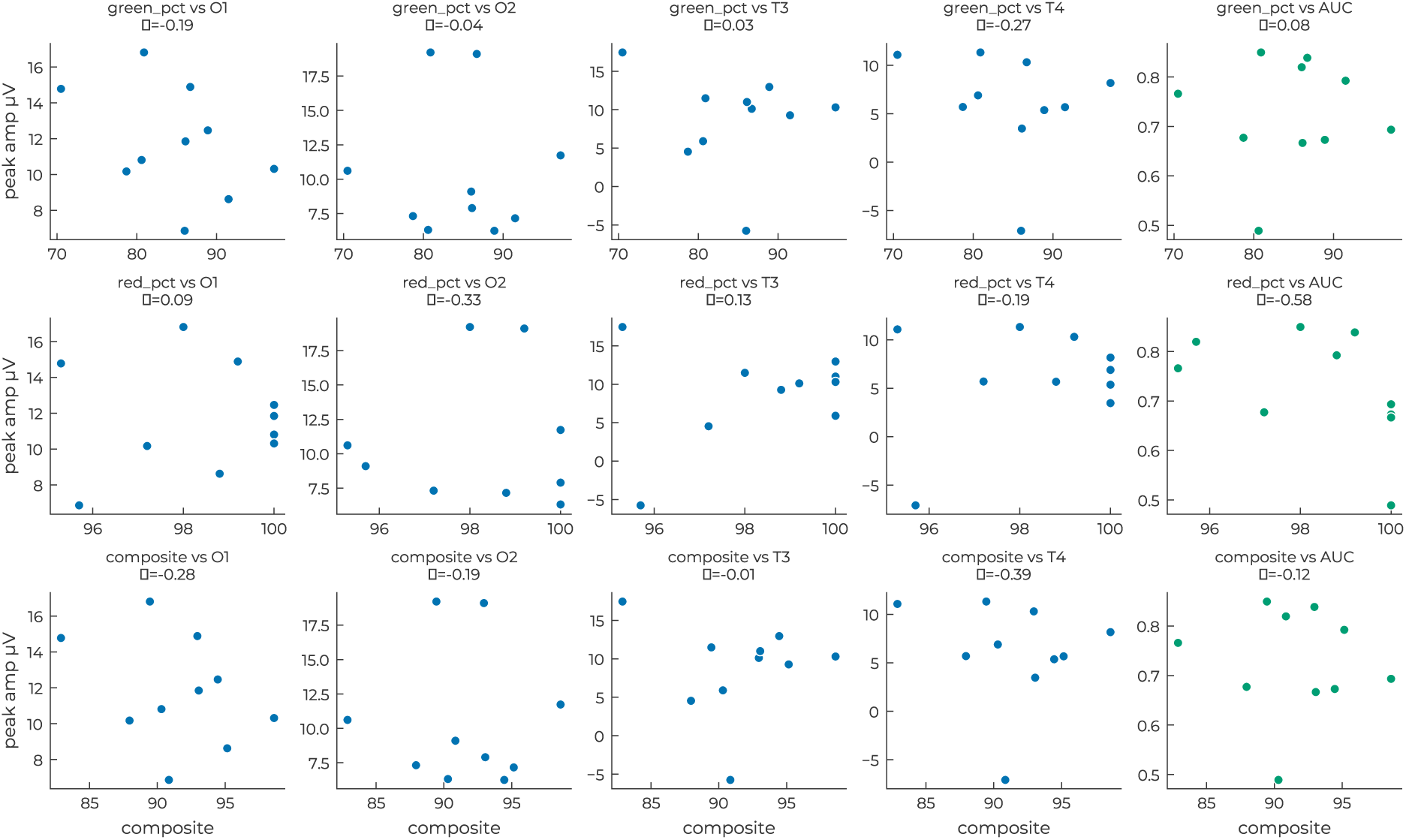
Scatter plots of per-subject behavioural scores (rows: green-gate rate, red-gate rate, composite) against per-subject difference-wave peak amplitude per channel and LDA AUC (columns). Spearman ρ for each pair is shown in the subplot title.

**Figure S12:**
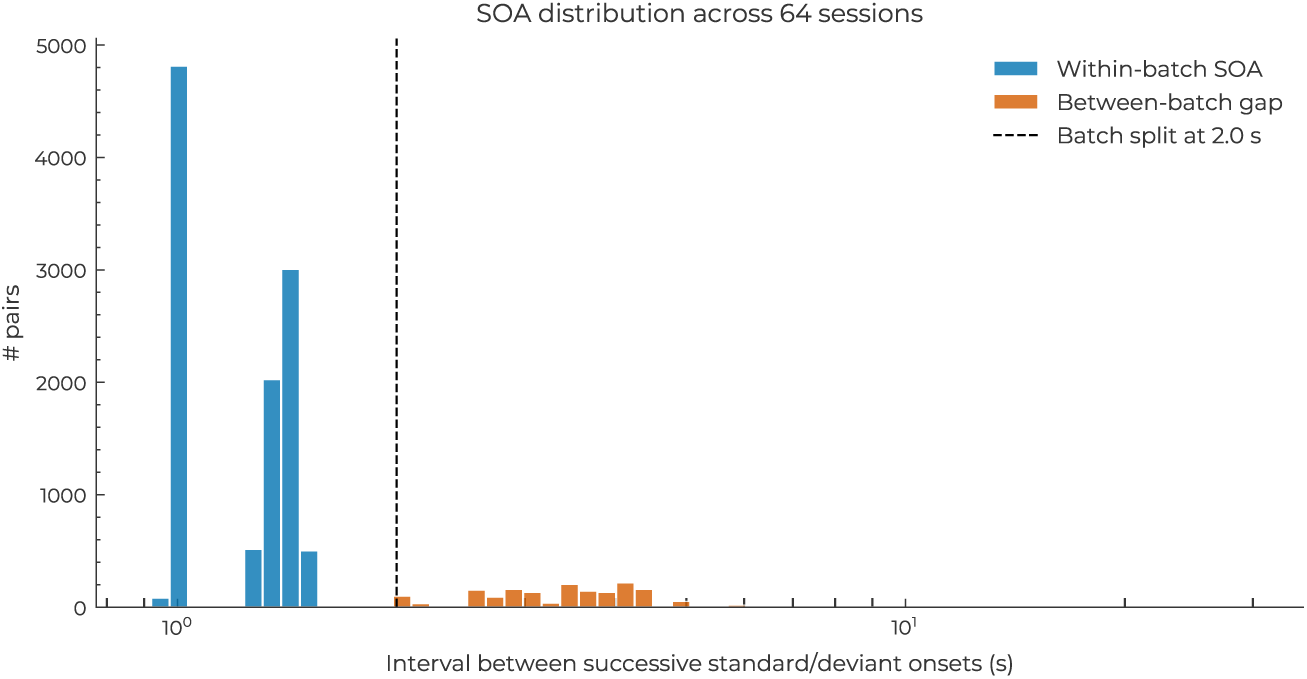
Distribution of stimulus onset asynchrony (SOA) across all game sessions.

**Figure S13:**
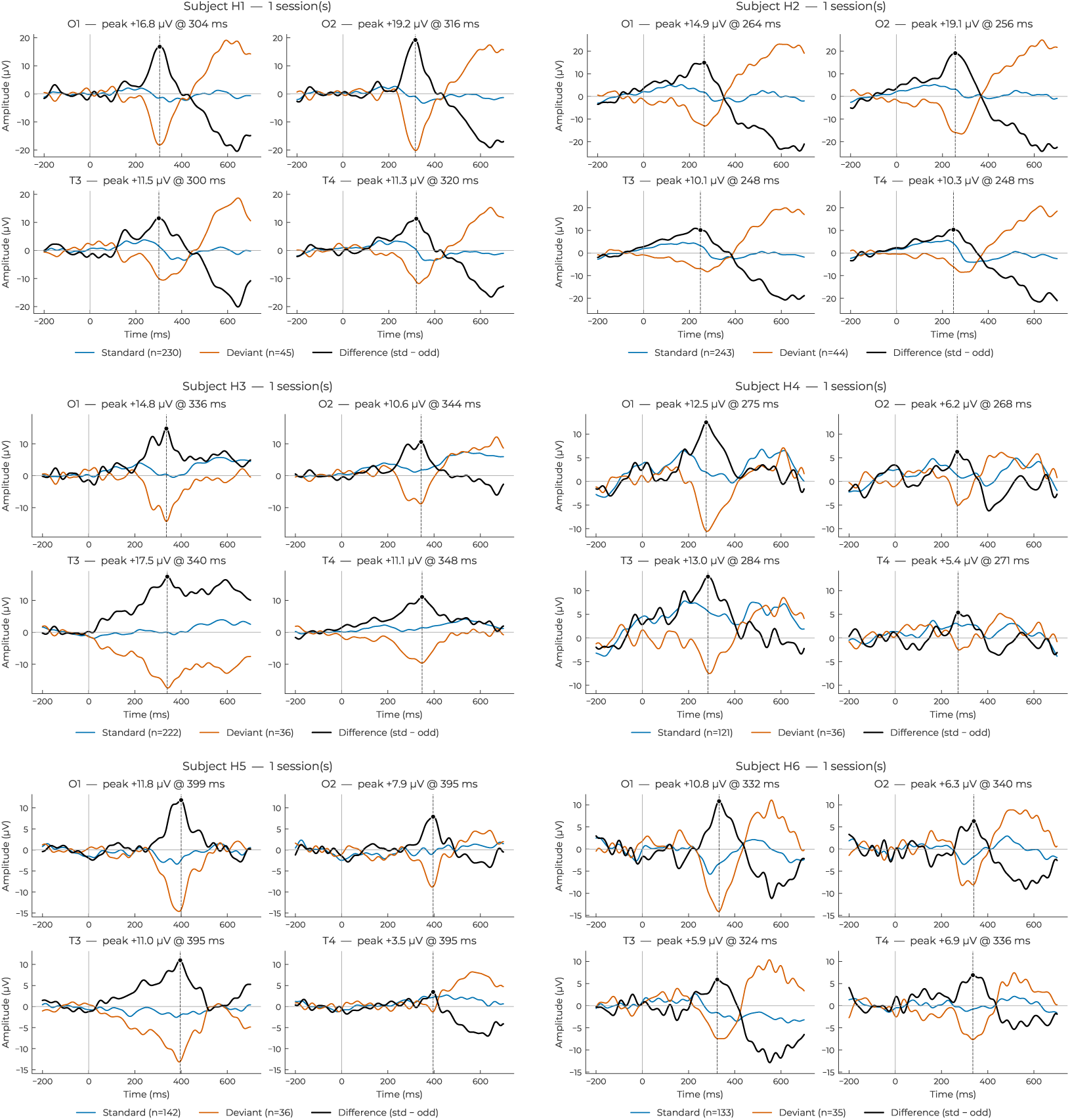
Individual subject ERP waveforms for healthy subjects H1–H6 (continued in fig-ure S14). Subjects sorted by O1 difference-wave peak amplitude (H1 strongest). Each panel shows one subject with four channels (O1, O2, T3, T4); standard (blue), deviant (orange), and difference (black, std − odd) waveforms are overlaid. Per-channel title shows the diff-wave peak amplitude and latency in 250–400 ms.

**Figure S14:**
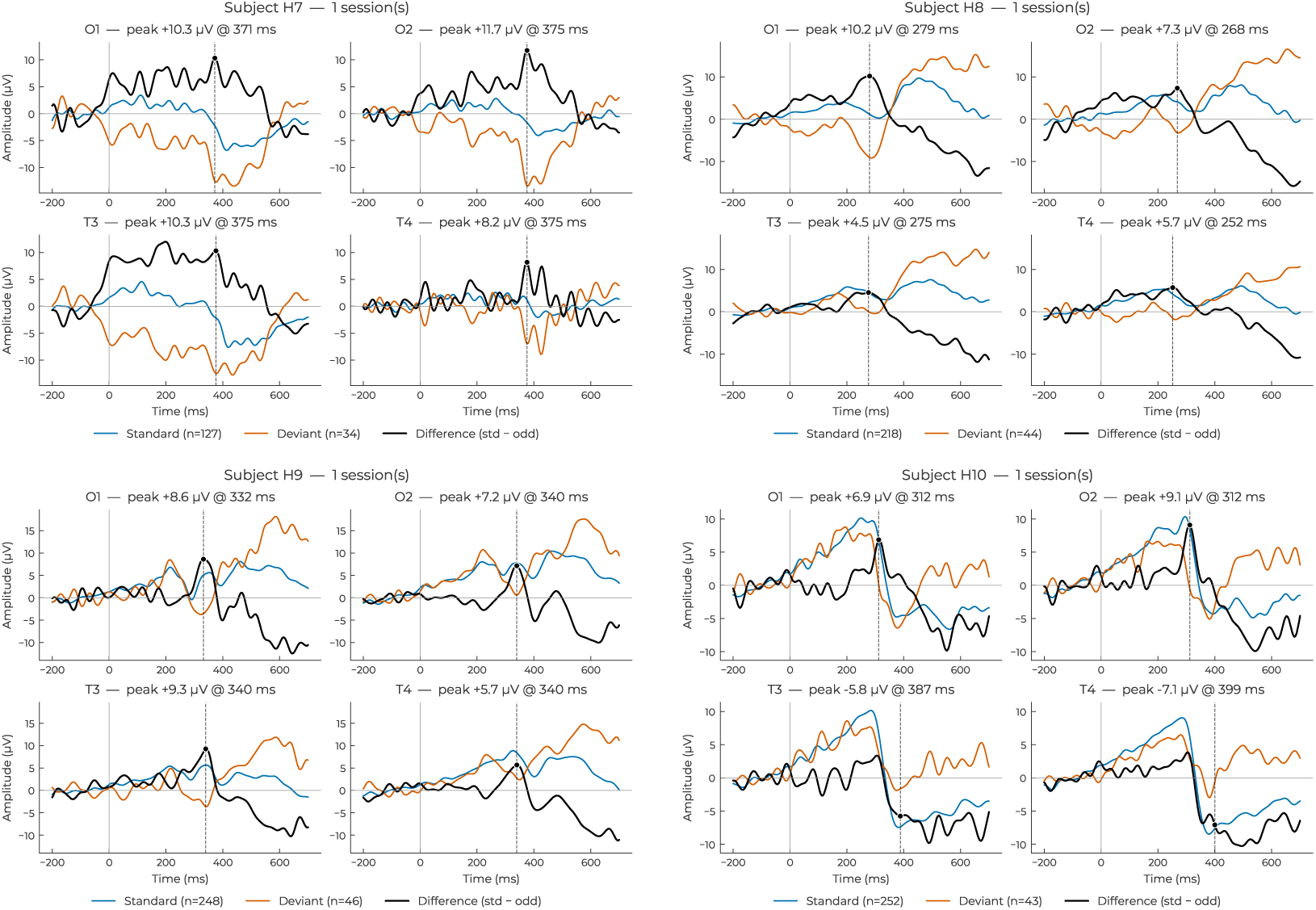
Individual subject ERP waveforms for healthy subjects H7–H10 (continued from figure S13). Same conventions.

**Figure S15:**
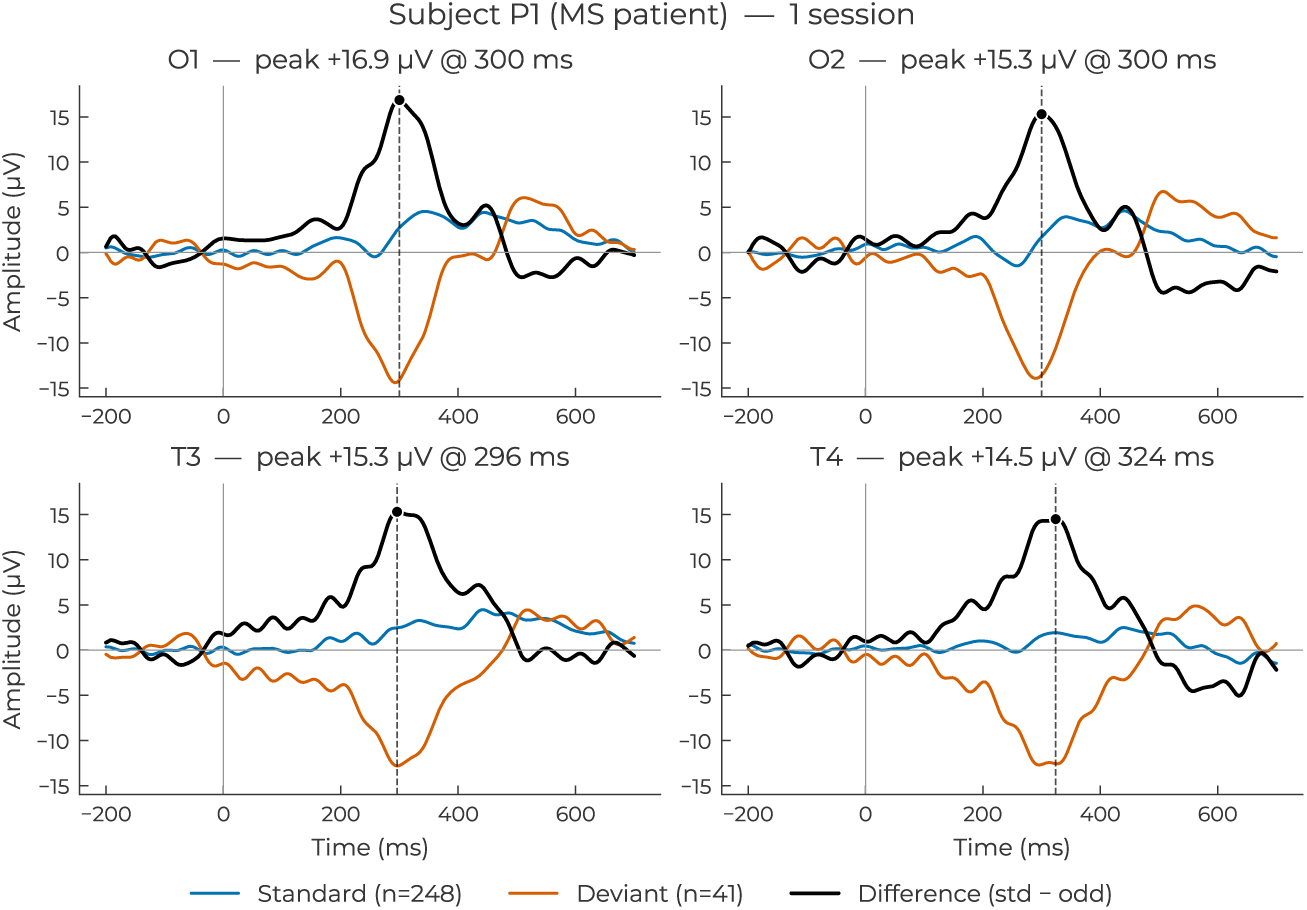
Individual subject ERP waveforms for the MS patient (P1), same conventions as figure S13.

## References

Gŕegoire Cattan, Anton Andreev, and Etienne Visinoni. Recommendations for integrating a p300-based brain–computer interface in virtual reality environments for gaming: An update. Computers, 9(4):92, 2020. doi: 10.3390/computers9040092.

Chun-Chuan Chen, Kai-Syun Syue, Kai-Chiun Li, and Shih-Ching Yeh. Neuronal correlates of a virtual-reality-based passive sensory p300 network. PLoS ONE, 9(11):e112228, 2014. doi: 10.1371/journal.pone.0112228.

Marco Congedo, Mathieu Goyat, Nicolas Tarrin, Gelu Ionescu, Ĺeo Varnet, Bertrand Rivet, Ronald Phlypo, Nisrine Jrad, Maria Acquadro, and Christian Jutten. “Brain Invaders”: a prototype of an open-source P300-based video game working with the OpenViBE platform. In 5th International Brain-Computer Interface Conference, Graz, Austria, 2011.

Stefan Debener, Reiner Emkes, Maarten De Vos, and Martin Bleichner. Unobtrusive ambulatory eeg using a smartphone and flexible printed electrodes around the ear. Scientific Reports, 5 (1):16743, 2015. doi: 10.1038/srep16743.

Reza Fazel-Rezai, Brendan Z. Allison, Christoph Guger, Eric W. Sellers, Sonja C. Kleih, and Andrea Kübler. P300 brain computer interface: current challenges and emerging trends. Frontiers in Neuroengineering, 5:14, 2012. doi: 10.3389/fneng.2012.00014.

Andrea Finke, Alexander Lenhardt, and Helge Ritter. The mindgame: A p300-based brain–computer interface game. Neural Networks, 22(9):1329–1333, 2009. doi: 10.1016/j.neunet.2009.07.003.

Ilya P. Ganin, Sergei L. Shishkin, and Alexander Y. Kaplan. A p300-based brain-computer interface with stimuli on moving objects: Four-session single-trial and triple-trial tests with a game-like task design. PLoS ONE, 8(10):e77755, 2013. doi: 10.1371/journal.pone.0077755.

Aitana Grasso-Cladera, Marko Bremer, Simon Ladouce, and Francisco Parada. A systematic review of mobile brain/body imaging studies using the p300 event-related potentials to investigate cognition beyond the laboratory. Cognitive, Affective, & Behavioral Neuroscience, 24 (4):631–659, 2024. doi: 10.3758/s13415-024-01190-z.

Christoph Guger, Gunther Krausz, Brendan Z. Allison, and Guenter Edlinger. Comparison of dry and gel based electrodes for p300 brain–computer interfaces. Frontiers in Neuroscience, 6, 2012. doi: 10.3389/fnins.2012.00060.

Vojislav Jovanović, Igor Petrušć, Vanja Ković, and Andrej M. Savić. The practical implications of re-referencing in erp studies: The case of n400 in the picture–word verification task. Diagnostics, 15(2):156, 2025. doi: 10.3390/diagnostics15020156.

Hussein Ghani Kaddoori. P300 event-related potentials in patients with multiple sclerosis. The Egyptian Journal of Neurology, Psychiatry and Neurosurgery, 59(1), 2023. doi: 10.1186/s41983-023-00726-3.

Jai Kalra, Prashasti Mittal, Nirmiti Mittal, Abhishek Arora, Utkarsh Tewari, Aviral Chharia, Rahul Upadhyay, Vinay Kumar, and Luca Longo. How visual stimuli evoked p300 is transforming the brain–computer interface landscape: A prisma compliant systematic review. IEEE Transactions on Neural Systems and Rehabilitation Engineering, 31:1429–1439, 2023. doi: 10.1109/TNSRE.2023.3246588.

Alexander Y. Kaplan, Sergei L. Shishkin, Ilya P. Ganin, Ivan A. Basyul, and Alexander Y. Zhigalov. Adapting the p300-based brain–computer interface for gaming: A review. IEEE Transactions on Computational Intelligence and AI in Games, 5(2):141–149, 2013. doi: 10.1109/TCIAIG.2012.2237517.

Ivo Kähner, Andrea Kübler, and Sebastian Halder. Rapid p300 brain-computer interface communication with a head-mounted display. Frontiers in Neuroscience, 9, 2015. doi: 10.3389/fnins.2015.00207.

Jongsu Kim and Sung-Phil Kim. A plug-and-play p300-based bci with calibration-free application. 2025. doi: 10.1101/2025.05.21.655021. Preprint.

Jonathan W. P. Kuziek, Abdel R. Tayem, Jennifer I. Burrell, Eden X. Redman, Jeff Murray, Jenna Reinen, Aldis Sipolins, and Kyle E. Mathewson. Real brains in virtual worlds: Validating a novel oddball paradigm in virtual reality. 2019. doi: 10.1101/749192. Preprint.

Seungchan Lee, Misung Kim, and Minkyu Ahn. Evaluation of consumer-grade wireless eeg systems for brain-computer interface applications. Biomedical Engineering Letters, 14(6): 1433–1443, 2024. doi: 10.1007/s13534-024-00416-w.

Man Li, Feng Li, Jiahui Pan, Dengyong Zhang, Suna Zhao, Jingcong Li, and Fei Wang. The mindgomoku: An online p300 bci game based on bayesian deep learning. Sensors, 21(5):1613, 2021. doi: 10.3390/s21051613.

D. Marshall, D. Coyle, S. Wilson, and M. Callaghan. Games, gameplay, and bci: The state of the art. IEEE Transactions on Computational Intelligence and AI in Games, 5(2):82–99, 2013. doi: 10.1109/TCIAIG.2013.2263555.

Leisi Pei, C. Shawn Green, and Guang Ouyang. Characterizing a highly excited and sustained brain response activity during gaming: P300-ce. Proceedings of the National Academy of Sciences, 122(30):e2502135122, 2025. doi: 10.1073/pnas.2502135122.

Igor Petrusic, Vojislav Jovanovic, Vanja Kovic, and Andrej M. Savic. P3 latency as a biomarker for the complexity of migraine with aura: Event-related potential study. Cephalalgia, 42(10): 1022–1030, 2022. doi: 10.1177/03331024221090204.

Terence W. Picton. The p300 wave of the human event-related potential. Journal of Clinical Neurophysiology, 9(4):456–479, 1992. doi: 10.1097/00004691-199210000-00002.

John Polich. Updating p300: An integrative theory of p3a and p3b. Clinical Neurophysiology, 118(10):2128–2148, 2007. doi: 10.1016/j.clinph.2007.04.019.

Andrej Savić, Romulus Lontis, Ning Jiang, Mirjana Popović, Dario Farina, Kim Dremstrup, and Natalie Mrachacz-Kersting. Movement related cortical potentials and sensory motor rhythms during self initiated and cued movements. In Biosystems & Biorobotics, pages 701–707. 2014. doi: 10.1007/978-3-319-08072-7_98.

Joanna E. M. Scanlon, Danielle L. Cormier, Kimberley A. Townsend, Jonathan W. P. Kuziek, and Kyle E. Mathewson. The ecological cocktail party: Measuring brain activity during an auditory oddball task with background noise. Psychophysiology, 56(11):e13435, 2019. doi: 10.1111/psyp.13435.

Mathias Sundgren, Vadim V. Nikulin, Liselotte Maurex, Åke Wahlin, Fredrik Piehl, and Tom Brismar. P300 amplitude and response speed relate to preserved cognitive function in relapsing–remitting multiple sclerosis. Clinical Neurophysiology, 126(4):689–697, 2015. doi: 10.1016/j.clinph.2014.07.024.

S. Sutton, M. Braren, J. Zubin, and E. R. John. Evoked-potential correlates of stimulus uncertainty. Science, 150(3700):1187–1188, 1965. doi: 10.1126/science.150.3700.1187.

Robin Vlieger, Duncan Austin, Deborah Apthorp, Elena Daskalaki, Artem Lensky, Dianne Walton-Sonda, Hanna Suominen, and Christian J. Lueck. The use of event-related potentials in the investigation of cognitive performance in people with multiple sclerosis: Systematic review. Brain Research, 1832:148827, 2024. doi: 10.1016/j.brainres.2024.148827.

Hiroki Watanabe and Yasushi Naruse. P300 as a neural indicator for setting levels of goal scores in educational gamification applications from the perspective of intrinsic motivation: An erp study. Frontiers in Neuroergonomics, 3:948080, 2022. doi: 10.3389/fnrgo.2022.948080.

